# Comparative analysis of 136,401 Admixed Americans and 419,228 Europeans reveals ancestry-specific genetic determinants of clonal haematopoiesis

**DOI:** 10.1101/2024.02.07.24302442

**Authors:** Sean Wen, Pablo Kuri-Morales, Fengyuan Hu, Abhishek Nag, Ioanna Tachmazidou, Sri Vishnu Vardhan Deevi, Haeyam Taiy, Katherine Smith, Douglas P. Loesch, Oliver S. Burren, Ryan S. Dhindsa, Sebastian Wasilewski, Jesus Alegre-Díaz, Jaime Berumen, Jonathan Emberson, Jason M. Torres, Rory Collins, Keren Carss, Quanli Wang, Slavé Petrovski, Roberto Tapia-Conyer, Margarete A. Fabre, Andrew R. Harper, George Vassiliou, Jonathan Mitchell

## Abstract

The development of clonal haematopoiesis (CH), the age-related expansion of mutated haematopoietic stem cell (HSC) clones, is influenced by genetic and non-genetic factors. To date, large-scale studies of CH have focused on individuals of European descent, such that the impact of genetic ancestry on CH development remains incompletely understood. Here, we investigate this by studying CH in 136,401 admixed participants from the Mexico City Prospective Study (MCPS) and 419,228 European participants from the UK Biobank (UKB). We observe that CH was significantly less common in MCPS compared to UKB (adjusted odds ratio (OR) = 0.56 [95% Cl = 0.55-0.59], *P* = 1.60 x 10^-206^), a difference that persisted when comparing MCPS participants whose genomes were >50% ancestrally Indigenous American to those whose genomes were >50% ancestrally European (adjusted OR = 0.76 [0.70-0.83], *P* = 1.78 x 10^-10^). Genome- and exome-wide association analyses in MCPS participants identified two novel loci associated with CH (*CSGALNACT1* and *DIAPH3*), and ancestry-specific variants in the *TCL1B* locus with opposing effect on *DNMT3A*-versus non-DNMT3A-CH. Meta-analysis of the MCPS and UKB cohorts identified another five novel loci associated with overall or gene specific CH, including polymorphisms at *PAPR11/CCND2*, *MEIS1* and *UBE2G1/SPNS3*. Our CH study, the largest in a non-European population to date, demonstrates the profound impact of ancestry on CH development and reveals the power of cross-ancestry comparisons to derive novel insights into CH pathogenesis and advance health equity amongst different human populations.

## Introduction

The overwhelming majority of genetic association studies to date have been performed on participants of European descent, most recruited from the United States and United Kingdom^1^. The study of non-European cohorts provides opportunities for novel discovery and orthogonal validation of risk variants/loci, and for improving our understanding of disease aetiology, whilst ensuring that the benefits of genomic research are broadly applicable^2^. This is especially true for the discovery of rare variants, as their origins are more recent and they tend to be more geographically clustered and population-specific^1,2^.

As with all human cells, haematopoietic stem cells (HSCs) accumulate somatic mutations with advancing age. Some of these mutations confer a fitness advantage to the affected HSC, promoting clonal expansion and engendering the phenomenon of clonal haematopoiesis (CH)^3–8^. CH is associated with an increased risk of haematological and other cancers^9^ and of non-oncological pathologies such as cardiovascular, renal, pulmonary and liver diseases^10–14^. Risk factors that contribute to increased CH include non-modifiable characteristics such as age and sex, exposures such as smoking or treatment with genotoxic drugs, and heritable genetic variation^15–17^. However, the impact of ancestry on CH development remains incompletely understood as to date studies of clonal haematopoiesis have focused largely on populations of European or European-dominant ancestry^15–17^.

To address this gap, we analysed whole-exome sequencing data to identify CH in 136,401 participants recruited to the Mexico City Prospective Study (MCPS). We compared the CH status in MCPS participants with 419,228 individuals from the United Kingdom Biobank (UKB), and performed intra-population analysis of MCPS individuals with varying proportions of European ancestry to delineate the contribution of ancestry to CH. We then performed single-variant genetic association analysis of common and rare germline variants, and gene-level genetic association analysis of rare germline variants with CH. The genetic association analyses were performed in MCPS individuals alone, and in a cross-ancestry meta-analysis combining MCPS and UKB participants. Our study, the largest investigation of CH in a non-European population to date, gives new insights into the contribution of ancestry to CH, discovers ancestry-specific risk variants, and identifies novel risk loci from combined analysis of UKB and MCPS individuals, among other findings.

## Results Prevalence of CH

Genetic ancestry at the continent level was determined for MCPS and UKB participants using *peddy*^18^ with the 1000 Genomes reference panel, and further analyses were restricted to 136,401 MCPS individuals with admixed Indigenous American, European, and African ancestry (≥95% *peddy*-predicted probability Admixed American) and 419,228 UKB individuals (≥95% *peddy*-predicted probability European, see Methods and Supplementary Tables 1 and 2). Somatic variants were called from whole exome sequencing data using MuTect2^19,20^ and filtered against a catalogue of predefined CH driver mutations in 15 genes, as described previously (see Methods and Supplementary Table 3)^21^. In total, we detected 4,656 somatic variants in 4,234 MCPS individuals in MCPS, and 22,518 somatic variants in 20,786 UKB individuals. The most recurrently mutated CH driver genes identified among MCPS participants were *DNMT3A, TET2*, *ASXL1*, *PPM1D*, and *TP53* (Fig. 1a), consistent with previous whole-exome sequencing studies of European-ancestry cohorts^15,16^. The prevalence of CH correlated strongly with age in both UKB and MCPS (*P*UKB and *P*MCPS < 2.2 x 10^-16^, Fig. 1b). The characteristics of CH variants identified displayed patterns in line with previous reports (Extended Data Fig. 1a-i, 2a-d), such as for mutation types (Extended Data Fig. 1f and g) and gene-specific CH associations with age (Extended Data Fig. 2a-d).

**Fig. 1.**
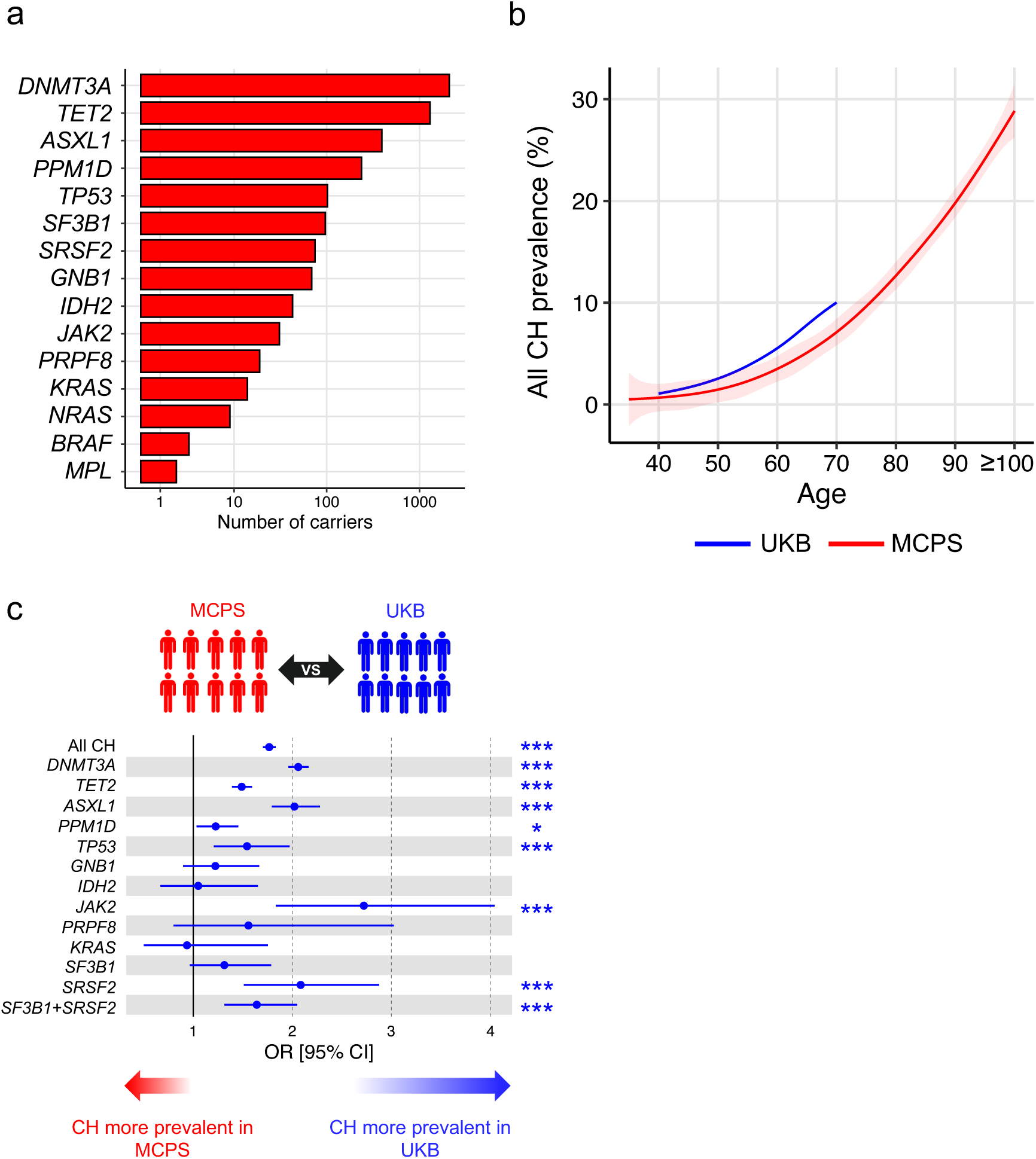
Prevalence of all CH in the MCPS and UKB. **a**, Number of individuals for each CH driver gene identified in MCPS. Driver genes ranked from the most to least number of individuals. **b**, Prevalence of CH by age. The centre line represents the fitted values from the general additive model with P-spline smooth class, and the shaded region represents the 95% confidence interval of the fitted values. MCPS individuals aged 100 or above were included as a single age group. **c**, Inter-population comparison of the prevalence of all CH and gene-specific CH in UKB compared to MCPS. Only CH driver genes identified in at least 10 individuals were included for analysis. Odds ratios and *P* values were derived from logistic regression model with all CH or gene-specific CH as outcome, and with age, sex, and smoking status included as covariates. *P* value *** < 0.001 ** < 0.01 * < 0.05.

Overall CH prevalence was significantly higher among UKB individuals (4.96%) compared to MCPS individuals (3.10%, *P*χ2 < 2.2 x 10^-16^), despite the MCPS cohort having an older average age than UKB (MCPS: range = 35-112, mean = 67.0; UKB: range = 40-70, mean = 60.8; Extended Data Fig. 2e). To account for the discrepancy in age distribution, we compared CH prevalence after age- and sex-matching, and found that CH prevalence amongst UKB individuals remained significantly higher than in MCPS individuals at all ages (overall CH prevalence 4.55% and 2.84%, respectively, *P*χ2 < 2.2 x 10^-16^, Extended Data Fig. 2f). Consistent with the increased prevalence of CH in UKB, we observed an increased risk of overall CH in UKB versus MCPS individuals in inter-population logistic regression with age, sex, and smoking included as covariates (OR = 1.77 [1.70-1.83], *P* = 1.60 x 10^-206^; Fig. 1c). We generally observed the same pattern when analysing CH driver genes individually, with significantly increased risk in UKB versus MCPS of CH driven by mutations in each of the 5 most common genes (*DNMT3A*, *TET2*, *ASXL1*, *PPM1D* and *TP53*), as well as *JAK2* and *SRSF2*. Focusing on the two most common CH genes, *DNMT3A* and *TET2*, the difference in magnitude of increased risk in UKB was particularly marked (OR = 2.06 [1.96-2.16], *P* = 1.52 x 10^-182^ and OR = 1.49 [1.39-1.59], *P* = 1.07 x 10^-30^, for *DNMT3A* and *TET2*, respectively). Importantly, whole-exome sequencing and variant calling was performed using the same workflow for the UKB and MCPS cohorts, resulting in similar variant allele frequency (VAF) and sequencing coverage profiles between cohorts across CH driver genes (Extended Data Fig. 3a-j). In a sensitivity analysis including sequencing coverage, in addition to age, sex, and smoking, as covariates in our logistic regression model led to similar results (Extended Data Fig. 4a). Furthermore, a similarly increased risk of CH among UKB individuals was observed when we matched individuals for age and sex, as an orthogonal approach to account for demographics differences between the two cohorts (Extended Data Fig. 4b-d and Supplementary Table 4).

## Association between ancestry and CH prevalence

To investigate the relationship between ancestry and CH further, we leveraged the admixed ancestry of the MCPS individuals^22^. The average proportion of Indigenous American, European, and African genome, as inferred by RFMix2.0^23^, across MCPS individuals included in our study was 66%, 31%, and 3%, respectively (Extended Data Fig. 5a). Their mosaic haplotype structure, incorporating multiple intercontinental genetic ancestries, provides an opportunity to robustly assess the relationship between genetic ancestry and CH prevalence. We found that the prevalence of CH was significantly higher in MCPS individuals whose genomes were >50% ancestrally European, compared to MCPS individuals whose genomes were >50% ancestrally Indigenous American across all age groups (Fig. 2a), with overall CH prevalence of 4.65% versus 2.82%, respectively (*P*χ2 < 2.2 x 10^-16^). Assessing genetic ancestry within MCPS as an ordinal variable further demonstrated a positive correlation between the fraction of individuals’ genomes derived from European ancestry with overall CH prevalence (Fig. 2b), even after adjusting for age, sex, and smoking (Extended Data Fig. 5b). Consistent with this, we observed that an increased risk of overall CH was associated with the proportion of European genome as a continuous variable in intra-population logistic regression with age, sex, and smoking included as covariates (beta = 0.85 [0.66-1.03], *P* = 6.60 x 10^-19^), and also with increased risk of *DNMT3A*-CH (beta = 1.15 [0.89-1.41], *P* = 4.74 x 10^-18^), *ASXL1*-CH (beta = 1.56 [0.97-2.14], *P* = 1.88 x 10^-7^), and *SRSF2*-CH (beta = 2.20 [0.86-3.54], *P* = 1.31 x 10^-3^) (Fig. 2c).

**Fig. 2.**
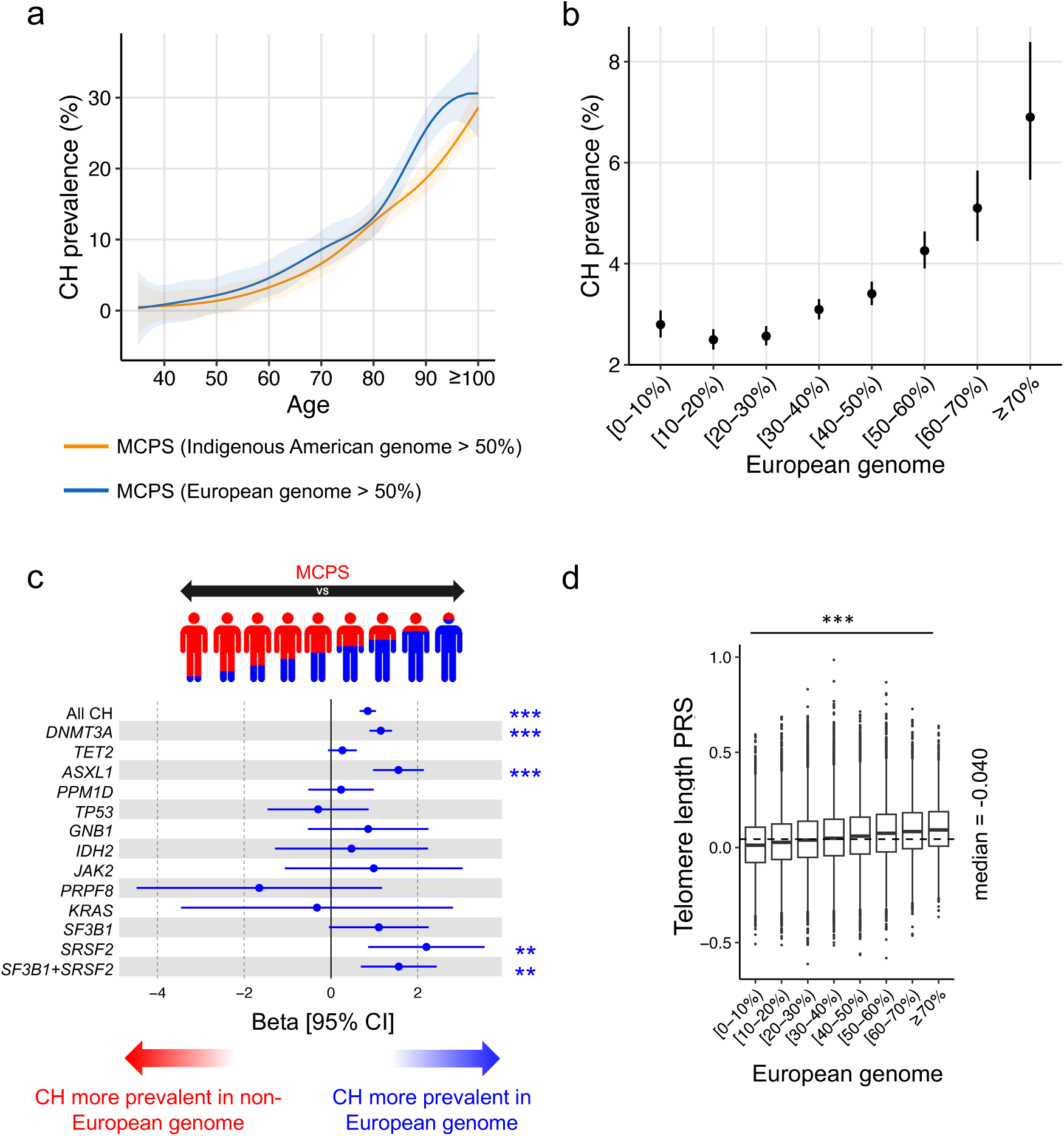
Ancestry association with CH prevalence in MCPS. **a**, Prevalence of all CH by age among individuals with >50% Indigenous American ancestry and individuals with >50% European ancestry. Ancestry genome proportion was inferred with RFMix2.0 software. The centre line represents the fitted values from the general additive model with P-spline smooth class, and the shaded region represents the 95% confidence interval of the fitted values. Individuals aged 100 or above were included as a single age group. **b**, Prevalence of all CH by proportion of European genome. Error bars represent 95% confidence interval. **c**, Intra-population comparison of the prevalence of all CH and gene-specific CH among individuals with varying degree of European and non-European (Indigenous American and African) genome. Only CH driver genes identified in at least 10 individuals were included for analysis. Beta coefficients and *P* values were derived from logistic regression model with all CH or gene-specific CH as outcome, and with age, sex, and smoking status included as covariates. **d**, Distribution of PRS-predicted telomere length across individuals with varying degree of European genome. PRS was constructed using telomere length-associated SNPs derived from GWAS analysis of 9,649 participants from MCPS with whole-genome sequencing-inferred telomere length available. *P* value was derived from the Kruskal-Wallis test. PRS, polygenic risk score. *P* value *** < 0.001 ** < 0.01 * < 0.05.

European ancestry of the haplotype block and gene locus in which the CH gene resides also positively correlated with prevalence of *DNMT3A*-, *SF3B1*-, and *SRSF2*-CH (Extended Data Fig. 5c and d), although this was largely explained by correlation between local and global (genome-wide) genetic ancestry (Supplementary Table 5). For example, association between *DNMT3A* haplotype ancestry and *DNMT3A*-CH was attenuated after including global ancestry in our model (beta = 0.22 [0.14-0.27], *P* = 4.09 x 10^-10^, and beta = 0.09 [0.02-0.17], *P* = 0.015, before and after including global ancestry as covariate). Therefore, while haplotype-and gene-level ancestry may contribute to CH risk, global ancestry plays a larger role in determining CH risk. Correspondingly, admixture mapping did not identify additional genomic intervals associated with CH at genome-wide significance (defined as *P* < 1.25 x 10^-6^).

Emerging evidence from European populations suggests that CH risk is influenced by telomere length^15,24–26^. Therefore, we performed telomere length GWAS among 9,649 MCPS individuals with WGS-derived telomere length inferred from TelSeq^27^ (Extended Data Fig. 6a), and used the leading SNPs from each of the 17 loci meeting the suggestive significance threshold (*P* < 5 x 10^-6^) in a polygenic risk score (PRS) model to calculate the genetically predicted telomere length across all 136,401 MCPS individuals (Extended Data Fig. 6b and Supplementary Table 6). We found that longer genetically predicted telomere length was associated with overall CH, as well as *DNMT3A*-, *TET2*, and *ASXL1*-CH, whereas shorter genetically predicted telomere length was associated with *PPM1D*-CH among MCPS individuals (Extended Data Fig. 6c and Supplementary Table 7). We also observed that MCPS individuals with higher proportions of European ancestry had longer genetically predicted telomere length based on our MCPS-derived PRS (Fig. 2d). This was also true when using a telomere length PRS derived from an ancestry-diverse population (TOPMed^28^) and an European population (UKB Europeans^29^, Extended Data Fig. 6d and e). For example, the telomere length PRS of MCPS individuals with ≥70% European genome was significantly greater compared to those with <10% European genome, based on PRS derived from MCPS (*P*Wilcoxon = 9.28 x 10^-74^). Notably, the association among MCPS individuals of increased European ancestry with CH was, in small part, attributed to longer telomere length (overall CH: beta = 0.85 [0.66-1.03] versus beta = 0.81 [0.63-1.00], *P*permuted < 0.01; *DNMT3A*-CH: beta = 1.15 [0.89-1.41] versus beta = 1.10 [0.84-1.36], *P*permuted < 0.01 for model without or with telomere length as covariate, respectively, Supplementary Table 8).

## Genome-wide common variant associations with CH

To further explore the contribution of common genetic variants (minor allele frequency (MAF) ý1%) to CH risk, we performed genome-wide association studies (GWAS) in MCPS individuals to evaluate both overall CH, and driver-gene specific CH, in addition to splicing factor CH (*SF3B1* and *SRSF2*) analysed in combination. Association analysis evaluating overall CH, including 4,234 cases and 132,167 controls, identified two genome-wide significant loci (*P* < 5 x 10^-8^ for lead variant) among MCPS individuals, namely the *TERT* and *TCL1B* loci (Fig. 3a, Table 1, Extended Data Fig. 7a). The *TERT* locus has shown the strongest association with CH in all European studies to date^15–17^. The lead *TERT* locus variant in our MCPS study was rs2853677, with the G allele (MAF = 23%) conferring an increased risk of overall CH (OR = 1.31 [1.24-1.37], *P* = 1.69 x 10^-24^), similar to what was reported in Europeans^15,16,30^. A conditional analysis at the *TERT* locus based on 22 previously reported significant variants^16^ at this locus did not identify additional novel independent variants (Supplementary Table 9).

**Fig. 3.**
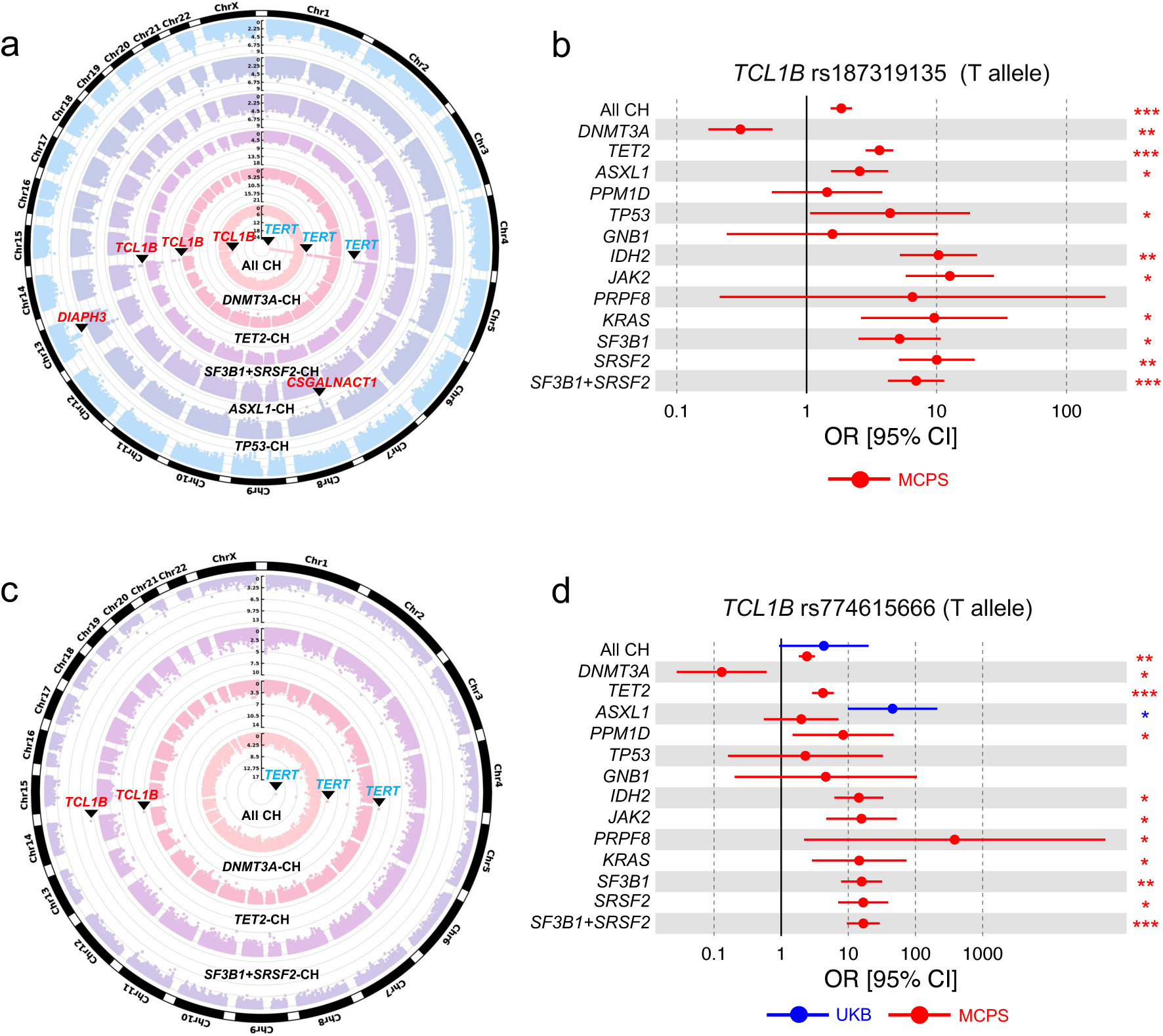
GWAS and ExWAS of all CH and gene-specific CH in MCPS. **a**, Circular Manhattan plot representing the common genetic variants (MAF ≥ 1%) included for GWAS for all CH and *DNMT3A*-, *TET2*-, *SF3B1+SRSF2*-, *ASXL1*-, and *TP53*-CH. *P* values on y-axis were derived from Firth logistic regression implemented by REGENIE software. Three new associations from MCPS (red) were identified as genome-wide significant (*P* value < 5 x 10^-8^) with the nearest gene of the leading SNP annotated for the respective locus. Previously reported associations from European populations indicated in blue. **b**, rs187319135 identified as genome-wide significant from overall CH, and *TET2*- and *SF3B1*+*SRSF2*-CH. Overall and gene-specific CH risk estimates conferred by the minor allele (T) in shown here for MCPS. Risk estimates shown when both CH or gene-specific CH cases and controls have minor allele count (MAC) ≥ 1. Risk estimates for UKB not included due to the absence of risk allele (MAC = 0) in CH individuals. **c**, Circular Manhattan plot representing the rare (MAF < 1%) and common genetic variants included for ExWAS for all CH and *DNMT3A*-, *TET2*-, and *SF3B1+SRSF2*-CH. *P* values on y-axis were derived from Firth logistic regression implemented by REGENIE software. Rare *TCL1B* promoter variant rs774615666 (red) was identified as exome-wide significant (*P* value < 1 x 10^-8^). Common *TERT* variant indicated in blue. **d**, rs774615666 identified as exome-wide significant from *TET2*- and *SF3B1*+*SRSF2*-CH. Overall and gene-specific CH risk estimates conferred by the minor allele (T) are shown here for MCPS and UKB. Risk estimates shown when both CH or gene-specific CH cases and controls have MAC ≥ 1. *P* values were derived from Firth logistic regression implemented by REGENIE software. *P* value *** < 5 x 10^-8^ (genome-wide significant), ** < 5 x 10^-6^ (suggestive), * < 0.05 (nominal).

**Table 1:**
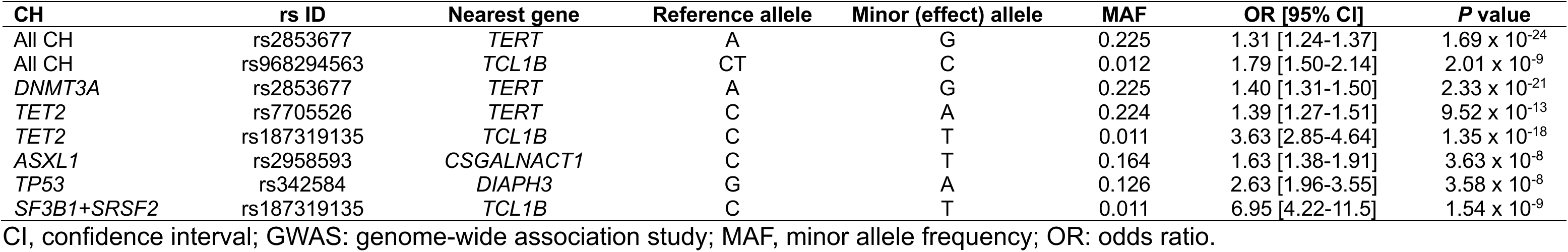
GWAS summary statistics of leading SNPs (smallest *P* value at each genome-wide significant locus) in MCPS. Related to Fig. 3a.

| CH | rs ID | Nearest gene | Reference allele | Minor (effect) allele | MAF | OR [95% CI] | *P* value |
| --- | --- | --- | --- | --- | --- | --- | --- |
| All CH | rs2853677 | *TERT* | A | G | 0.225 | 1.31 [1.24-1.37] | $1.69 \times 10^{-24}$ |
| All CH | rs968294563 | *TCL1B* | CT | C | 0.012 | 1.79 [1.50-2.14] | $2.01 \times 10^{-9}$ |
| *DNMT3A* | rs2853677 | *TERT* | A | G | 0.225 | 1.40 [1.31-1.50] | $2.33 \times 10^{-21}$ |
| *TET2* | rs7705526 | *TERT* | C | A | 0.224 | 1.39 [1.27-1.51] | $9.52 \times 10^{-13}$ |
| *TET2* | rs187319135 | *TCL1B* | C | T | 0.011 | 3.63 [2.85-4.64] | $1.35 \times 10^{-18}$ |
| *ASXL1* | rs2958593 | *CSGALNACT1* | C | T | 0.164 | 1.63 [1.38-1.91] | $3.63 \times 10^{-8}$ |
| *TP53* | rs342584 | *DIAPH3* | G | A | 0.126 | 2.63 [1.96-3.55] | $3.58 \times 10^{-8}$ |
| *SF3B1+SRSF2* | rs187319135 | *TCL1B* | C | T | 0.011 | 6.95 [4.22-11.5] | $1.54 \times 10^{-9}$ |

CI, confidence interval; GWAS: genome-wide association study; MAF, minor allele frequency; OR: odds ratio.

In addition to replicating the well-established *TERT* locus association, we also discovered two novel low-frequency risk variants located 82.9kb (rs968294563, MAF = 1.24%) and 115kb (rs187319135, MAF = 1.06%) upstream of *TCL1B* (T-cell leukaemia/lymphoma protein 1B) on chromosome 14. These were in strong linkage disequilibrium (D’ > 0.99, r^2^ = 0.86) and conditional analysis did not reveal independent signals. In addition to an increased risk of overall CH (OR: 1.85 [1.53-2.23], *P* = 2.69 x 10^-9^), rs187319135 (T allele) also displayed genome-wide significant associations with *TET2*- and *SF3B1*+*SRSF2*-CH (OR = 3.63 [2.85-4.64], *P* = 1.35 x 10^-18^ and OR = 6.95 [4.22-11.5], *P* = 1.54 x 10^-9^, respectively, Extended Data Fig. 7b and c). At the nominally significant threshold (*P* < 0.05), it was also associated with increased risk of seven other gene-specific CH subtypes, but a decreased risk of *DNMT3A*-CH (OR = 0.31 [0.18-0.55], *P* = 1.30 x 10^-6^, Fig. 3b, Supplementary Table 10).

Notably*, TCL1B* is located adjacent to *TCL1A* and whilst CH-associated risk variants (rs10131341 and rs2887399) have been previously identified at the *TCL1B-TCL1A* locus in European population analyses^15,31^, rs187319135 is highly enriched in

Indigenous American (Mexican) versus European ancestry (*MCPS Variant Browser* https://rgc-mcps.regeneron.com/home/ (2023): MAF = 1.62% and MAF = 0.06%, respectively). Although rs187319135 was in high linkage disequilibrium with rs10131341 (D’ = 0.95, r^2^ = 0.024), specifically the minor allele of rs187319135 was observed to be often co-inherited alongside the major allele of rs10131341 (Extended Data Fig. 8a), it remained associated with overall CH, and *TET2-* and *SF3B1+SRSF2*-CH at genome-wide significance threshold even after conditioning on rs10131341 (Extended Data Fig. 8b and c). We also replicated the association between rs10131341 and *DNMT3A*-, *TET2*, and *ASXL1*-CH at nominal significance threshold (*P* < 0.05), and the association remained significant after conditioning on rs187319135 (Extended Data Fig. 8d and e). Taken together, our observations suggest independence of these signals.

In addition to the *TCL1B*-*TCL1A* locus, additional novel loci were identified through GWAS of *ASXL1*- and *TP53*-CH in MCPS (Table 1). The lead variant associated with *ASXL1*-CH was rs2958593 at the *CSGALNACT1* locus (Extended Data Fig. 7d), and the T allele (MAF = 16%) conferred an increased risk exclusively of *ASXL1*-CH (OR = 1.63 [1.38-1.91], *P* = 3.63 x 10^-8^, Extended Data Fig. 9a). The lead variant associated with *TP53*-CH was rs342584 at the *DIAPH3* locus (Extended Data Fig. 7e), where the A allele (MAF = 13%) conferred an increased risk exclusively of *TP53*-CH (OR = 2.63 [1.96-3.55], *P* = 3.58 x 10^-8^, Extended Data Fig. 9b). Conditioning on the lead SNP at each locus did not identify additional independent risk variants.

Many of the reported lead variant associations with CH in European populations were replicated among MCPS individuals. Among previously reported lead CH variants^15–17,32^, 7 of 22 (32%) for overall CH, 12 of 20 (60%) for *DNMT3A*-, 5 of 7 (71%) for *TET2*-, 2 of 3 (67%) for *ASXL1*-, and 1 of 3 (33%) for *JAK2*-CH were nominally significant (*P* < 0.05) among MCPS individuals (Extended Data Fig. 10 and 11a-d, Supplementary Table 11).

## Exome-wide rare variant associations with CH

We next sought to identify rare germline variants associated with CH in the MCPS cohort using exome-wide association analysis^33^. Common risk variants identified from GWAS were largely recapitulated in our whole-exome association analysis at exome-wide significance (*P* < 1 x 10^-8^ Supplementary Table 12). Focusing on rare variants (MAF < 1%), we identified a rare variant in the *TCL1B* promoter (rs774615666, MAF = 0.33%), that increased risk for *TET2*-CH (OR = 4.19 [2.89-6.06], *P* = 2.77 x 10^-10^) and *SF31B+SRSF2*-CH (OR = 16.7 [9.53-29.4], *P* = 9.15 x 10^-13^, Fig. 3c, Table 2, and Extended Data Fig. 12a-b). Notably, rs774615666 was associated, at nominal or suggestive significance thresholds (*P* < 0.05 and *P* < 1 x 10^-6^, respectively), with increased risk of *PPM1D*-, *IDH2*-, *JAK2*-, *PRPF8*-, *KRAS*, *SF3B1*- and *SRSF2*- CH, but a decreased risk of *DNMT3A*-CH (Fig. 3d, Supplementary Table 13). Within the MCPS cohort, rs774615666 was unique to Indigenous American (Mexican) ancestry (*MCPS Variant Browser* https://rgc-mcps.regeneron.com/home/ (2023): MAF = 0.50%, versus MAF = 0% for European ancestry), and was still significant after conditioning on the previously identified *TCL1A* upstream (rs10131341) and promoter (rs2887399) variants for *TET2*-CH (OR = 4.15 [2.77-6.21], *P* = 5.97 x 10^-9^) and *SF3B1+SRSR2*-CH (OR = 15.8 [7.50-29.2], *P* = 7.37 x 10^-9^). Notably, rs774615666 was in high linkage disequilibrium with the GWAS identified rs187319135 (D’ = 0.95, r^2^ = 0.32), yet rs187319135 remained associated with *TET2*-CH at genome-wide significance threshold after conditioning on rs774615666 while there were residual associations with overall CH and gene-specific CH (*DNMT3A*-, *ASXL1*-, *IDH2*-, *JAK2*-, *SRSF2*-, and *SF3B1+SRSF2*-CH) at suggestive or nominal significance thresholds (*P* < 0.05; Extended Data Fig. 13a and b). Conversely, rs774615666 remained associated with overall CH and gene-specific CH (*TET2*-, *SF3B1*-, *SRSF2*-, and *SF3B1+SRSF2*-CH) at nominal significance threshold (*P* < 0.05) after conditioning on rs187319135 (Extended Data Fig. 13c and d).

**Table 2:** ExWAS summary statistics of leading SNPs (smallest *P* value at each exome-wide significant locus) in MCPS. Related to Fig. 3c.

| CH | rs ID | Nearest gene | Reference allele | Minor (effect) allele | MAF | OR [95% CI] | *P* value |
| --- | --- | --- | --- | --- | --- | --- | --- |
| *TET2* | rs774615666 | *TCL1B* | C | T | 0.0032 | 4.19 [2.89-6.06] | 2.77 x 10<sup>-10</sup> |
| *SF3B1+SRSF2* | rs774615666 | *TCL1B* | C | T | 0.0032 | 16.7 [9.53-29.4] | 9.15 x 10<sup>-13</sup> |

CI, confidence interval; ExWAS: exome-wide association study; MAF, minor allele frequency; OR, odds ratio.

To further delineate the overall CH and gene-specific CH risks conferred by these variants, we stratified individuals into those with rs187319135 risk allele only, rs774615666 risk allele only, and both rs187319135 and rs774615666 risk alleles and compared them with individuals with no risk alleles for both rs187319135 and rs774615666 (Extended Data Fig. 14a). We observed individuals carrying only the rs187319135 risk allele to be at higher risk of overall CH and *TET2*-, *ASXL1*-, *TP53*-, *IDH2*-, *JAK2*-, *KRAS*-, *SRSF2*-, and *SF3B1+SRSF2*-CH, but decreased risk to *DNMT3A*-CH (*P* < 0.05, Extended Data Fig. 14b). We further observed individuals carrying only the rs774615666 risk allele to be at increased risk of *TET2*-, *SF3B1*-, *SRSF2*-, and *SRSF2-SF3B1*-CH (*P* < 0.05). Taken together, the ancestry-specific variants rs187319135 and rs774615666 may be partly independent of one another, and functional assays, such as Hi-C, may help elucidate the causal variant(s).

## Cross-ancestry meta-analysis of CH

Cross-ancestry GWAS meta-analysis across UKB and MCPS individuals, yielded five novel CH-associated loci, one for overall CH and four for driver gene-specific CH (Fig. 4a and Supplementary Table 14). In total, 15 loci were detected for overall CH, including a novel association in the *GACAT3*-*CYRIA* locus (rs7566405, A allele: OR = 0.94 [0.93-0.96], *P* = 1.03 x 10^-8^). Driver-gene specific CH GWAS for *DNMT3A* revealed 19 loci, of which two were novel: *MEIS1* (rs2280334, T allele: OR = 1.08 [1.05-1.10], *P* = 2.30 x 10^-8^) *and PARP11-CCND2* (rs582975, T allele: OR = 0.93 [0.91-0.95], *P* = 1.26 x 10^-8^; Fig. 4b)*. MEIS1* is a gene with critical roles in both normal and malignant hematopoiesis^34^, while *CCND2* is recurrently mutated in acute myeloid leukaemia (AML)^35^. PARP11 plays a role in inflammatory responses by regulating IFN-I signalling^36^. In *TET2*-CH, five loci were detected including a novel association within the *UBE2G1-SPNS3* locus (rs73332852, T allele; OR = 1.50 [1.33-1.69], *P* = 6.69 x 10^-11^, Fig. 4c). For *ASXL1*-CH, two loci were identified including the novel *PPIA-H2AZ2*-*PURB* locus (rs13245012, T allele: OR = 1.20 [1.13-1.27], *P* = 1.60 x 10^-9^, Fig. 4d). One additional novel locus (*PDXDC1*) was discovered from our UKB-only GWAS for *DNMT3A*-CH (rs28545123, C allele; OR = 0.90 [0.87-0.93], *P* = 2.15 x 10^-8^).

**Fig. 4.**
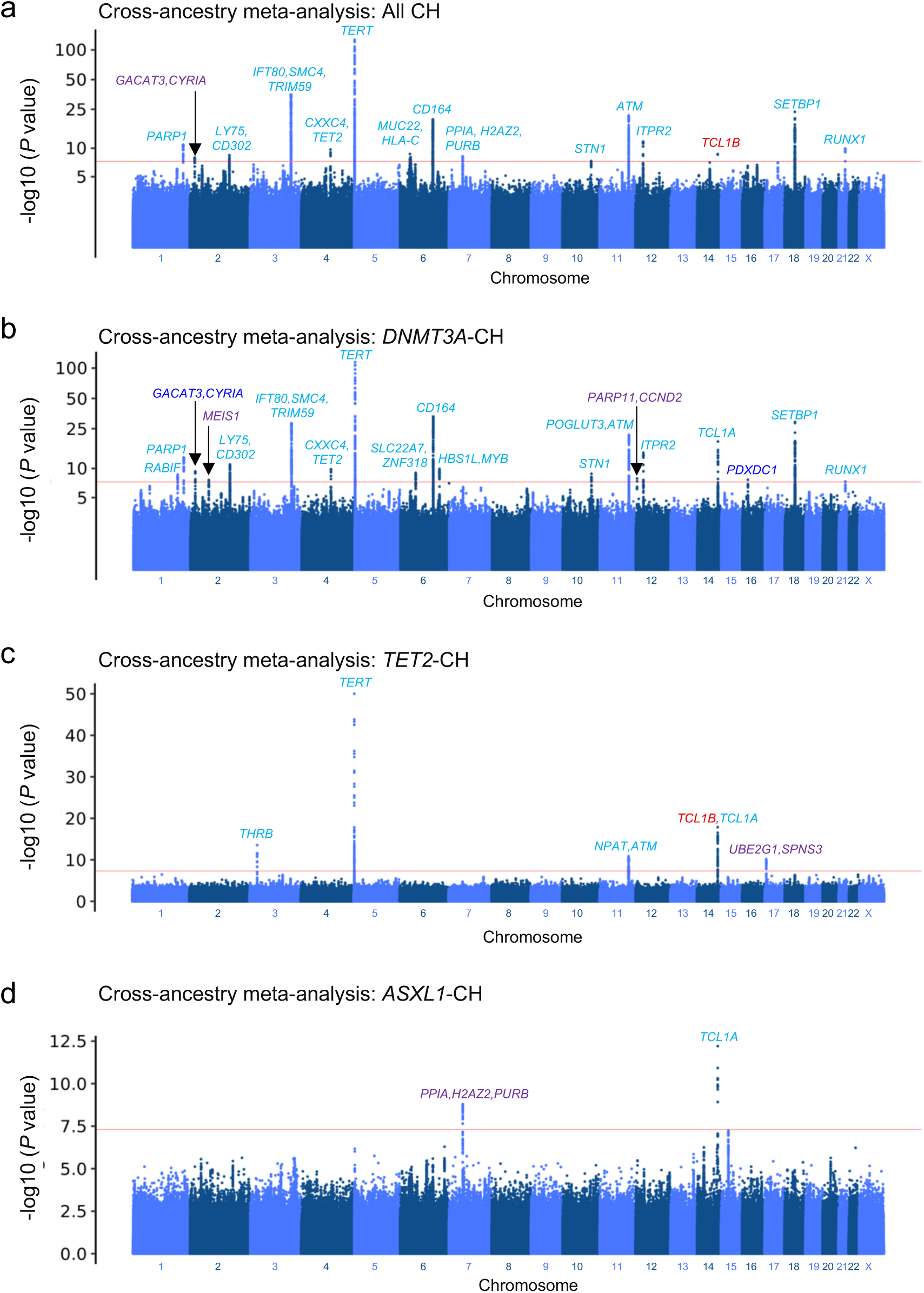
Cross-ancestry GWAS meta-analysis of all CH and gene-specific CH in MCPS and UKB. **a-d**, Manhattan plot representing the common genetic variants (MAF ≥ 1%) included for GWAS in MCPS and UKB Europeans for all CH **(a)**, and *DNMT3A*-**(b)**, *TET2*-(**c**), and *ASXL1*-**(d)** CH. *P* values on y-axis were derived from inverse variance-weighted average method implemented by METAL software. Five novel loci from meta-analysis of MCPS and UKB (purple) were identified as genome-wide significant (*P* value < 5 x 10^-8^) with the nearest gene of the leading SNP annotated for the respective locus. Previously reported associations from European populations indicated in light blue, novel association from European population identified in our study indicated in dark blue, and novel associations from cross-ancestry meta-analysis identified in our study indicated in purple.

As the rare variants found to be associated with CH were ancestry-specific, it was not surprising that cross-ancestry exome-wide association meta-analysis did not identify additional risk variants. Utilising a gene-level collapsing test, which aggregates all qualifying rare germline variants for a given gene, may increase statistical power by testing the combined effect of rare variants^33,37^. As in previous work^33^, we used eleven different qualifying variant (QV) models to maximise discovery across potential genetic architectures (see Methods). While gene-level collapsing analysis in the MCPS cohort did not identify any genes significantly associated with CH, the MCPS-UKB meta-analysis replicated the previously reported *CHEK2* association with overall CH (flexible-damaging QV model: OR = 1.64 [1.44-1.86], *P* = 3.95 x 10^-13^) and with *DNMT3A*-CH (flexible-damaging QV model: OR = 1.78 [1.51-2.08], *P* = 3.71 x 10^-11^, Extended Data Fig. 15a, c, and e, and Supplementary Table 15). Scrutiny of the qualifying rare variants in *CHEK2* revealed that the majority were ancestry-specific (Extended Data Fig. 15b, d, and f and Supplementary Table 16). Notably, *CHEK2* c.1100del constituted 72% of all *CHEK2* qualifying variants in UKB individuals, but only 4.7% in MCPS individuals. Taken together, cross-ancestry meta-analysis yielded several novel CH susceptibility loci driven by common variants.

## Discussion

We present the largest analysis of CH in a non-European population to date and leverage genetic ancestry to uncover fundamental novel insights into the aetiology of this common age-related phenomenon. We demonstrate that the prevalence of CH among 136,401 individuals from Latin America (MCPS cohort) is 1.6-fold lower than the prevalence among almost 419,228 individuals in UKB (3.10% versus 4.96%, respectively), with our CH prevalence in Europeans consistent with previously studied European-majority populations of a similar age distribution (range: 4.3-6%)^15–17^. The implications of population-level differences in CH prevalence include considerations around population-wide or targeted CH screening planning and strategy, resource allocation, appropriateness of CH screening as part of routine health screening and risk assessment for CH-related diseases, such as myeloid neoplasms^9,38^ and cardiovascular or other pathologies.

Prior to our study, non-European populations, including those from Admixed Americans, were underrepresented in population-based studies, which precluded robust estimation of CH prevalence, including at the driver-gene specific level. Indeed, previous studies have yielded discrepant results: for example, one large study reported CH prevalence to be similar across different ancestries^16^, whilst smaller but more ancestry-diverse studies suggested a lower prevalence among self-reported Latinos or Hispanics compared to Europeans^4,17,39^. Utilising MCPS, currently the largest population cohort of genetically-defined Admixed Americans, we demonstrate that the odds ratio of CH among MCPS participants compared to UKB participants was 0.56. The difference was particularly striking for certain CH driver genes, with odd ratios of 0.49 for *DNMT3A*-CH and 0.37 for *JAK2*-CH in MCPS compared to UKB, for example. It is tenable that the substantial lower prevalence of *JAK2*-CH underlies the lower prevalence of myeloproliferative neoplasms, most of which are *JAK2*-driven, in Hispanic populations^40,41^.

Differences in CH prevalence between populations from different continents may be influenced by lifestyle, environmental, and socioeconomic factors, as well as by genetic ancestry. We leveraged the admixed genetic nature of MCPS participants, with individual genomes comprising different proportions of European versus Indigenous American ancestry, to isolate the effect of genetic ancestry on CH while mitigating the potential confounding impact of environmental differences between populations. We demonstrated within MCPS that increased risk of CH is associated with a higher fraction of European compared to Indigenous American ancestry, revealing that genetic ancestry is an important determinant of CH risk.

Genetic association analyses of CH among European-majority populations have identified 51 risk loci for overall or gene-specific CH^15–17,32^. Here, common germline variant association analysis of CH among MCPS individuals identified two novel risk loci, and ancestry specific genetic variants in the *TCL1B* locus. Leading variants rs2958593 and rs342584, located within *CSGALNACT1* and *DIAPH3* respectively, were associated exclusively with *ASXL1*- and *TP53*-CH, respectively. Additionally, leading variant rs187319135, located upstream of *TCL1B,* was associated with opposing effects on *DNMT3A*-CH compared to non-DNMT3A-CH. This variant was independent of the *TCL1A* upstream variant (rs10131341) previously identified in European studies^15,16^. Also, cross-ancestry meta-analysis of UKB and MCPS individuals identified 5 novel loci associated with overall or gene specific CH (*GACAT3/CYRIA, MEIS1, PARP11/CCND2, UBE2G1/SPNS3, PPIA/H2AZ2/PURB*). Together with the 2 novel loci (*CSGALNACT1* and *DIAPH3*) identified in the MCPS-only GWAS and 1 novel locus (*PDXDC1*) identified in the UKB-only GWAS, we increase the number of germline associations with CH from 51 to 59.

Our exome-wide association study identified a rare *TCL1B* promoter risk variant (rs774615666) associated with *TET2*- and *SF3B1+SRSF2*-CH at exome-wide significance, in LD with the more common *TCL1B* GWAS variants (rs187319135 and rs968294563). Both ExWAS- and GWAS-identified risk variants were similarly associated with opposing effects on *DNMT3A*-CH compared to non-DNMT3A-CH. rs187319135 remained associated with CH, in particular *TET2*-CH, after conditioning on rs774615666, suggesting that rs187319135 is partly independent of rs774615666. It is also noteworthy that rs774615666 was also persistently associated with *TET2*- and *SF3B1+SRSF2*-CH, albeit not at exome-wide significance threshold, after conditioning on rs187319135. Further fine-mapping and experimental approaches such as Hi-C may be warranted to further understand the functional role and any potential interactions between these risk variants.

It is notable that rare variants are relatively recent evolutionary occurrences and tend to be ancestry-specific^42^. Indeed, the *TCL1B* upstream (rs187319135) and *TCL1B* promoter risk variant (rs774615666) were less common in Indigenous American compared to European ancestry (*MCPS Variant Browser* https://rgc-mcps.regeneron.com/home/ (2023): rs187319135 MAF = 1.6% and 0.06%, respectively; rs774615666 MAF = 0.5% and 0%, respectively). Whereas the CH-associated *CHEK2* c.1100delC risk variant was less common in Indigenous American compared to European ancestry (*MCPS Variant Browser* https://rgc-mcps.regeneron.com/home/ (2023): MAF = 0% and 0.2%, respectively), consistent with previous reports^43,44^. In comparison, 49% of previously identified common risk loci associated with CH in Europeans were successfully replicated among MCPS participants in our study. This is consistent with evolutionary theory suggesting that common variants are relatively old, many originating prior to human migration out of Africa, and therefore prevalent in many populations^1,43,44^.

One method of assessing the functional impact of common or rare variants identified from genome-or exome-wide association analysis, respectively, is expression quantitative trait loci (eQTL) analysis. For example, while the previously reported *TCL1A* upstream variant (rs10131341) was not associated with *TCL1A* expression, the T allele of the *TCL1A* promoter variant (rs2887399) has been shown to be associated with decreased *TCL1A* expression in the Genotype-Tissue Expression (GTEx) project^45^, and was subsequently validated experimentally^31^. The five novel variants identified in our study were significantly associated with the expression of nearby genes in one or more human tissues (rs2958593 with *CSGALNACT1*, rs342584 with *DIAPH3,* rs2280334 with *MEIS1-AS3,* rs73332852 with *CYB5D2,* and rs13245012 with *H2AZ2* and *ZMIZ2*)^45^. However, eQTL analysis of the *TCL1B* promoter risk variant was hampered by the lack of non-European-specific variants in publicly available European-majority eQTL datasets. Indeed, the *TCL1B* upstream or promoter risk variants are not catalogued in the GTEx project^45^. While there are emerging non-European eQTL resources, these datasets are relatively small and preclude robust assessment of low frequency genetic variants^46^. This highlights the urgency of establishing large-scale non-European population-based resources, including eQTL databases, to allow equitable research in diverse ancestries.

Collectively, we identify substantial differences in the prevalence of CH and its subtypes between populations and discover ancestry-specific genetic determinants. Our work demonstrates that the investigation of CH in diverse populations can reveal fundamental biological insights, with differential implications for population-level precision medicine, and ultimately the advancement of global health equality.

## Methods

### Study population

Participants were included from two population-based studies, namely the Mexico City Prospective Study (MCPS) and United Kingdom Biobank (UKB).

MCPS is a prospective cohort of more than 150,000 adults, aged at least 35 years, and recruited between 1998 and 2004 from the contiguous urban districts of Coyoacán and Iztapalapa in Mexico City^22,47^. Of these, 141,046 individuals were whole exome sequenced. UKB is a prospective cohort of approximately 500,000 adults, aged between 40 to 70 years, and recruited since 2007^48,49^, 469,809 of which were whole exome sequenced.

The MCPS study was approved by the Mexican Ministry of Health, the Mexican National Council for Science and Technology, and the University of Oxford, and the UKB study has approval from the North-West Multi-centre Research Ethics Committee (11/NW/0382).

### Whole-exome sequencing pre-processing, variant calling, and CH detection

Whole-exome sequencing data for genomic DNA from MCPS and UKB was generated by Regeneron Genetics Centre as previously described^22,48,50^. Sequencing libraries were generated using IDT xGen v1 capture kit, sequenced on the NovaSeq6000 platform in 75-bp paired-end mode, and subsequently demultiplexed using the 10-bp index barcodes to obtain the sequencing reads for each sample in FASTQ format.

The FASTQ files were processed at AstraZeneca as previously described^21^. Sequencing read alignment to the GRCh38 genome reference and germline variant detection, for exome-wide association analysis and gene-level collapsing analysis, were performed using the Illumina DRAGEN Bio-IT Platform Germline Pipeline v3.0.7. Somatic variant calling was performed using GATK MuTect2^19,20^. A panel of normals was created from 200 of the youngest UKB participants without a haematologic malignancy diagnosis to remove potential recurrent artifacts with GATK *FilterMutectCalls*. The *--orientation-bias-artifact-priors* option was also specified to remove read orientation artifacts based on priors generated with *LearnReadOrientationModel*.

In both MCPS and UKB, samples were selected on the basis of (1) contamination <4% computed by VerifyBAMID software^51^, (2) gender concordant between clinically reported and chromosome X:Y consensus coding sequence (CCDS) coverage ratios, (3) ≥94.15% of CCDS r22 bases^52^ covered with ≥10x coverage, (4) within 4SDs of mean genetic principal components 1-4 as computed by the *peddy* software^18^, and (5) SNP array QC (genotype missingness ≤10%). Samples from MCPS were additionally selected based on within 2 standard deviations (SDs) of the mean read-depth distribution, no pairs with kinship >0.45, and probability ≥0.95 of Admixed American ancestry. Samples from UKB were additionally selected based on no pairs with kinship >0.1769 and probability ≥0.95 of European ancestry. Kinship and and ancestry were inferred using the KING and *peddy* softwares, respectively^18,53,54^. *peddy* Admixed American and European ancestry probabilities were computed with the 1000 Genomes Admixed American and European reference panel, respectively. Post-QC, 136,401 individuals from MCPS and 419,228 individuals from UKB were included in our study (Supplementary Tables 1 and 2).

CH somatic mutations were identified from a predefined list in a panel of 74 genes known to be recurrently mutated in myeloid neoplasm^10,17^. Mutations were further restricted to those with 3-40% variant allele frequency (VAF), mutant allele supported by ≥3 reads, and variant site with ý10x coverage. Fifteen CH genes which demonstrated increased prevalence of driver variants with age were retained and included in our analyses^21^.

### Telomere length polygenic risk score

GWAS of telomere length was performed on 9,649 MCPS participants with whole-genome sequencing (WGS) available^22^. Specifically, telomere length was inferred from WGS using TelSeq^27^, and REGENIE^55^ was used for genetic association analysis with age, sex and first ten genetic principal components included as covariates as described above. Leading SNPs from 17 loci meeting the suggestive significance threshold (*P* < 5 x 10^-6^) were identified and were subsequently used to compute the telomere length polygenic risk scores (PRS) for all 136,401 MCPS participants. No evidence of genomic inflation was observed (inflation factor = 1.03). Additional telomere length PRSs were computed using telomere length-associated variants previously identified from Europeans from UKB^29^ (https://github.com/siddhartha-kar/clonal-hematopoiesis/blob/main/mendelian_randomization/tl.txt) and Hispanics/Latinos from Trans-Omics for Precision Medicine (TOPMED; Table 1 of Taub *et al*.)^28^. PRSs were computed using allelic dosages in PLINK 2 with the *cols=scoresums* option to obtain the raw (non-averaged) values^56^.

### Local ancestry inference, intra-population analysis, and admixture mapping

Proportions of European, Indigenous American, and African ancestry were inferred at each genomic interval (window) using the RFMix2.0 software as previously described^22,23^. In total, 39,861 genomic intervals were defined by RFMix2.0. Association between European ancestry and CH prevalence among MCPS participants (intra-population) was assessed at 3 levels (global, haplotype, and gene) using logistic regression with continental ancestry fraction as the exposure and CH as the outcome. For a given CH driver gene, the haplotype and gene levels were defined using the genomic intervals overlapping with the haplotype and genomic region, respectively, in which the CH driver gene resides. Therefore, the gene level interval(s) is contained within the haplotype level intervals. The haplotype and gene level ancestry estimates were averaged across the two alleles for each individual and categorised into European homozygous, European heterozygous, and non-European (America or Africa) homozygous based on European ancestry thresholds of ≥95%, 45-55%, and ≤5%, respectively. The association between European ancestry at the haplotype or gene level and gene-specific CH prevalence was assessed using an additive model whereby the non-European homozygous was the reference group. Age, sex, and smoking status were included as covariates and logistic regression was performed using the *glm* function as implemented by the *stats* package in R (v4.2.2).

Furthermore, the beta coefficients were assessed from the association between global European ancestry and CH before versus after MCPS-derived telomere length PRS was included as covariate in the logistic regression model. To this end, we applied a permutation test whereby the CH case and control labels were shuffled prior to fitting the model with or without the telomere length PRS as covariate. The differences in beta coefficient from the model with versus without the telomere length PRS was then computed (Δβ_permuted_). This process was repeated 100 times to establish the null distribution. Then, the non-permuted (observed) differences in beta coefficient from the model with versus without the telomere length PRS was computed (Δβ_observed_). Finally, the *P* value was derived by computing the percentage of times | Δβ_permuted_| > |Δβ_observed_|. Significant change in the beta coefficient after versus before telomere length PRS was included into the model was defined with *P* < 0.05.

Admixture mapping was performed by assessing the association between all genomic windows with overall CH and gene-specific CH. Genomic windows with *P* value < 1.25 x 10^-6^ (0.05/39,861 genomic windows) was considered as genome-wide significant. Age, sex, smoking status, and global European ancestry were included as covariates and logistic regression was performed using the *glm* function as implemented by the *stats* package in R (v4.2.2).

### Genetic association analyses

For MCPS, genome-and exome-wide association analysis (GWAS and ExWAS, respectively) of germline variants with overall CH and gene-specific CH was performed using the REGENIE software^55^. Briefly, the individuals were genotyped using Illumina Global Screening Array v.2 beadchip as previously described^22^. Sample-level QC was performed to remove samples with genotype missingness > 10% and related samples. Variant-level QC was performed to remove non-autosomal variants, variants with missingness ≥ 2%, variants with C>G, G>C, A>T, or T>A base change, variants in long-range LD regions, and insertions or deletions, and to retain variants with MAF ≥ 1%, with Hardy-Weinberg Equilibrium (HWE) *P* < 10^-6^, and variants pruned for LD r2 < 0.1 (windows of 50 SNPs and a size step of 5 SNPs). The remaining variants were used in step 1 of REGENIE. In step 1 of REGENIE, whole genome regression model was fitted using genotyped variants to each CH phenotype, and a set of genomic leave-one-chromosome-out (LOCO) predictions were returned. The model was fitted separately for each of the 22 autosomes and chromosome X. In step 2 of REGENIE, the germline variants (imputed with TOPMED reference panel)^22^ and WES-identified variants^37^ for GWAS and ExWAS, respectively, were tested for association with each CH phenotype using Firth logistic regression based on the additive model. Age, sex, and the first ten genetic principal components were included as covariates in both steps of REGENIE while the LOCO predictions were additionally included as covariate in step 2 of REGENIE. The summary statistics from each chromosome were then combined prior to downstream analysis, and restricted to variants with imputation quality score > 0.6, and minor allele count (MAC) of ≥5 in cases and controls^28^. A filter of MAF ≥ 1% was additionally applied for variants identified from GWAS^15^. Statistically significant GWAS and ExWAS variants were defined with *P* < 5 x 10^-8^ and *P* < 1 x 10^-8^, and the leading SNP for each locus was defined with the SNP with the smallest *P* value^15,28,37^. A locus was considered novel when there was no previously reported CH-associated genome-wide significant SNPs falling within ±1Mb of the leading SNP. For UKB, GWAS was performed using the REGENIE software as described above for MCPS. Briefly, individuals were genotyped using UK Biobank Axiom Array (UKBB) and UKB BiLEVE Axiom Array (UKBL). Sample-level QC was performed to retain individuals of European descent, and to remove samples with genotype missingness > 5%, samples with non-XX or -XY chromosome configuration, and samples with high heterozygosity. Variant-level QC was perform to remove variants with missingness ≥ 2%, non-autosomal variants, variants with C>G, G>C, A>T, or T>A base change, variants in long-range LD regions or in commonly inverted regions, insertions or deletions, and variants with different allele frequencies between UKBB and UKBL (defined with P < 10-12 when Fisher exact test applied on genotype counts), and to retain variants with MAF ≥ 1%, variants with HWE *P* < 10^-6^, and variants pruned for LD r2 < 0.1 (windows of 50 SNPs and a size step of 5 SNPs). The remaining samples and variants were used in step 1 of REGENIE. Variants imputed with Haplotype Reference Consortium (HRC) panel, and additionally with UK10K + 1000 Genomes panel for variants not present in HRC^49^, were used in step 2 of REGENIE. The variant coordinates were additionally converted from GRCh37 to GRCh38 human reference genome assembly with CrossMap^57^. ExWAS was performed on WES-identified variants using Fisher’s exact two-sided test based on the allelic model as previously described^33,37^.

Gene-level collapsing analysis for MCPS and UKB was performed using Fisher’s exact two-sided test as previously described^33,37^. Eleven different sets of qualifying variant models were assessed on the basis of variant effect on protein-coding sequence, MAF from gnomAD and internal test cohort, Rare Exome Variant Ensemble Learner (REVEL) score^58^, and Missense Tolerance Ratio (MTR)^59^. The QV models were “syn” (synonymous, MAF ≤0.005%), “flexdmg” (non-synonymous, MAF_global_ ≤0.05%, MAF_any given ancestry_ ≤0.1%, REVEL score ≥0.25), “flexnonsynmtr” (non-synonymous, MAF_global_ ≤0.05%, MAF_any given ancestry_ ≤0.1%, MTR <0.78 or MTR centile <50), “UR” (non-synonymous, MAFglobal = 0%, REVEL score ≥0.25), “URmtr” (“UR” and MTR <0.78 or MTR centile <50), “raredmg” (missense, MAF_global_ ≤0.005%, REVEL score ≥0.25), “raredmgmtr” (“raredmg” and MTR <0.78 or MTR centile <50), “ptv” (protein-truncating, MAF_global_ ≤0.1%, MAF_any given ancestry_ ≤0.1%), “ptv5pcnt” (protein-truncating, MAF_global_ ≤5%, MAF_any given ancestry_ ≤5%), “ptvraredmg” (“ptv” and “raredmg”), and, “rec” (non-synonymous, MAF_global_ ≤0.5%, MAF_any given ancestry_ ≤0.5%). All qualifying variants were assessed in a dominant model, with the exception of “rec”, which was assessed in a recessive model. Statistically significant genes were defined by a study-wide significance threshold, *P* < 1 x 10^-8^ ^37^.

Genetic association analyses were repeated with the case and control groups randomly assigned to each individual to obtain the permuted *P* values for genomic inflation assessment. MCPS and UKB did not display high levels of genomic inflation with overall CH: inflation factor was 1.02 and 1.12 for GWAS, 1.01 and 1.02 for ExWAS, and 1.01 and 1.03 for gene-level collapsing analysis.

### Genetic association meta-analyses

Meta-analysis across MCPS and UKB was performed using cross-ancestry (stratified) meta-analysis based on the summary statistics returned from GWAS, ExWAS, and gene-level collapsing analyses^2^. GWAS meta-analysis was performed using the inverse variance-weighted average (IVW)-based method by summarising the effect size of each variant from both cohorts. GWAS meta-analysis was additionally performed using the sample size-based method implemented in METAL^60^, which is more robust than IVW when combining summary statistics with large effect size standard errors. Statistically significant variants were defined with *P* < 5 x 10^-8^ from both IVW-and sample size-based methods. ExWAS meta-analysis was performed using the sample size-based method, and statistically significant variants were defined with *P* < 1 x 10^-8^. Both IVW-and sample size-based methods were implemented by the METAL software^60^. Gene-level collapsing meta-analysis was performed using Cochran-Mantel-Haenszel (CMH) test using the *mantelhaen.test* function as implemented by the *stats* package in R (v4.2.2).

## Code availability

All analyses was performed using publicly available software and web-based applications as indicated in the Methods section.

## Competing interests

S.W., F.H., A.N., I.T., S.V.V.D., H.T., K.S., K.C., D.P.L., O.S.B., R.S.D., S.W., Q.W., S.P., M.A.F., A.R.H., J.M. are current employees and/or stockholders of AstraZeneca. R.C. is the chair of the data monitoring committee of the PROMINENT trial and the deputy chair of a not-for-profit clinical trial company (PROTAS) unrelated to this work.

## Data Availability

Summary statistics for GWAS, ExWAS, gene burden association analysis, baseline phenotype association analysis, and mortality analysis are provided in the Supplementary Tables. Individual-level UK Biobank data may be requested via applicable to the UK Biobank. Individual-level MCPS data may be requested via Data and Sample Access Policy available on the Oxford-hosted webpage (http://www.ctsu.ox.ac.uk/research/mcps).

## Notes

### Funding Statement

The generation of the UKB data was funded by the UKB Exome Sequencing Consortium (UKB-ESC) members: AbbVie, Alnylam Pharmaceuticals, AstraZeneca, Biogen, Bristol-Myers Squibb, Pfizer, Regeneron, and Takeda. The MCPS has received funding from the Mexican Health Ministry, the National Council of Science and Technology for Mexico, the Wellcome Trust (058299/Z/99), Cancer Research UK, British Heart Foundation, and the UK Medical Research Council (MC_UU_00017/2). The generation of the MCPS data was funded through an academic partnership among the National Autonomous University of Mexico, the University of Oxford, Regeneron, AstraZeneca, and Abbvie. These funding sources had no role in the design, conduct, or analysis of the study or the decision to submit the manuscript for publication.

