## Supplemental Figures for "Comparative analysis of 136,401 Admixed Americans and 419,228 Europeans reveals ancestry-specific genetic determinants of clonal haematopoiesis"

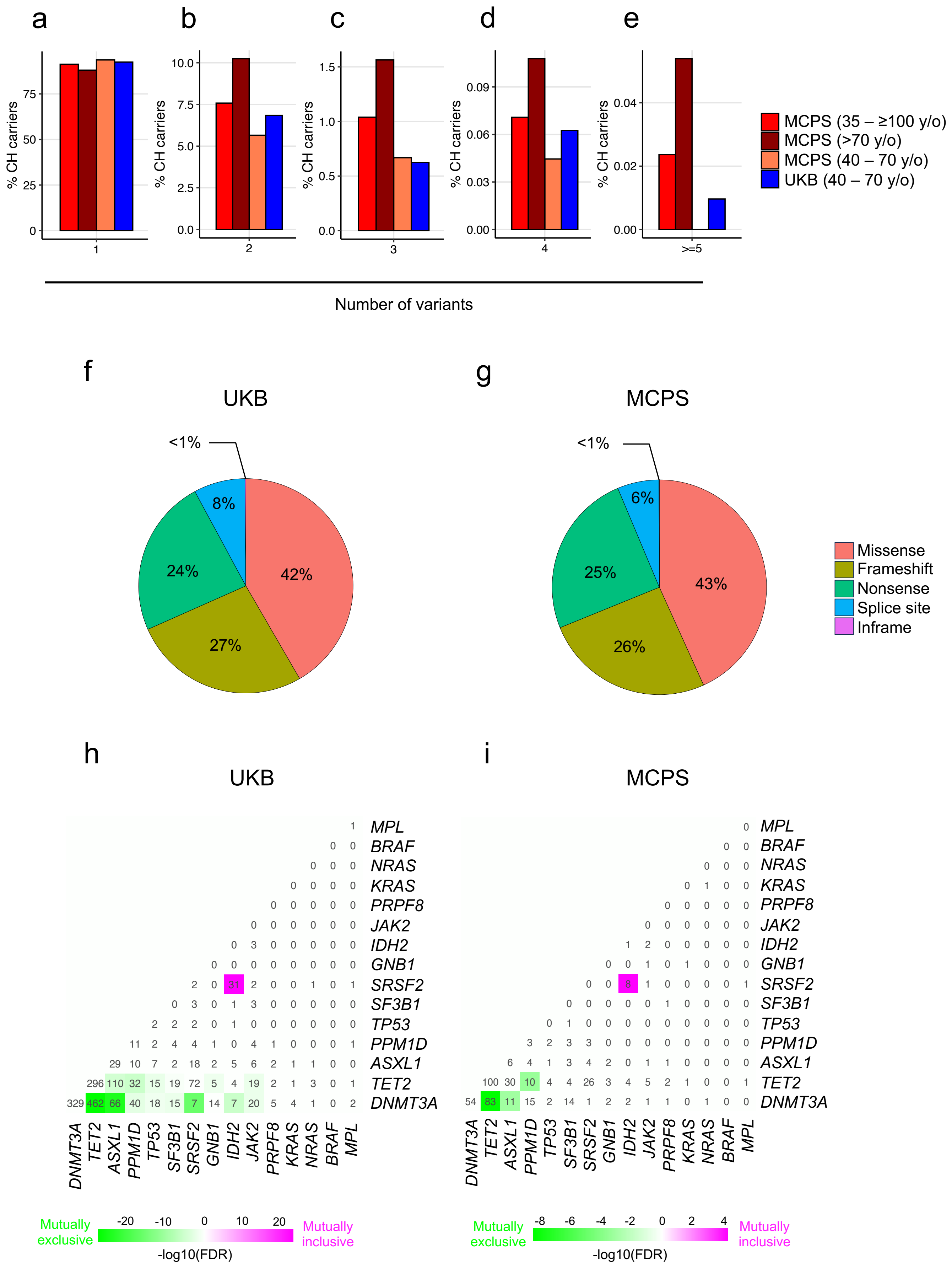

a
b
c
d
e
% CH carriers
MCPS (35 – ≥100 y/o)
MCPS (>70 y/o)
MCPS (40 – 70 y/o)
UKB (40 – 70 y/o)
1
2
3
4
>=5
Number of variants
f
UKB
<1%
8%
24%
42%
27%
g
MCPS
<1%
6%
25%
43%
26%
Missense
Frameshift
Nonsense
Splice site
Inframe
h
UKB
i
MCPS
MPL
BRAF
NRAS
KRAS
PRPF8
JAK2
IDH2
GNB1
SRSF2
SF3B1
TP53
PPM1D
ASXL1
TET2
DNMT3A
Mutually exclusive
Mutually inclusive
-log10(FDR)

**Extended Data Fig. 1 | Prevalence and characteristics of all CH. a-e**, Percentage of all CH individuals with 1 (**a**), 2 (**b**), 3 (**c**), 4 (**d**), or more than 5 (**e**) CH driver gene variants stratified by different age groups. **f,g**, CH driver gene variants stratified by consequence on protein-coding sequence in UKB (**f**) and MCPS (**g**). **h,i**, Assessment of co-occurrence or mutual exclusivity of CH driver genes among individuals with at least two mutated CH driver genes in UKB (**h**) and MCPS (**i**). ORs and *P* values were derived from logistic regression model.

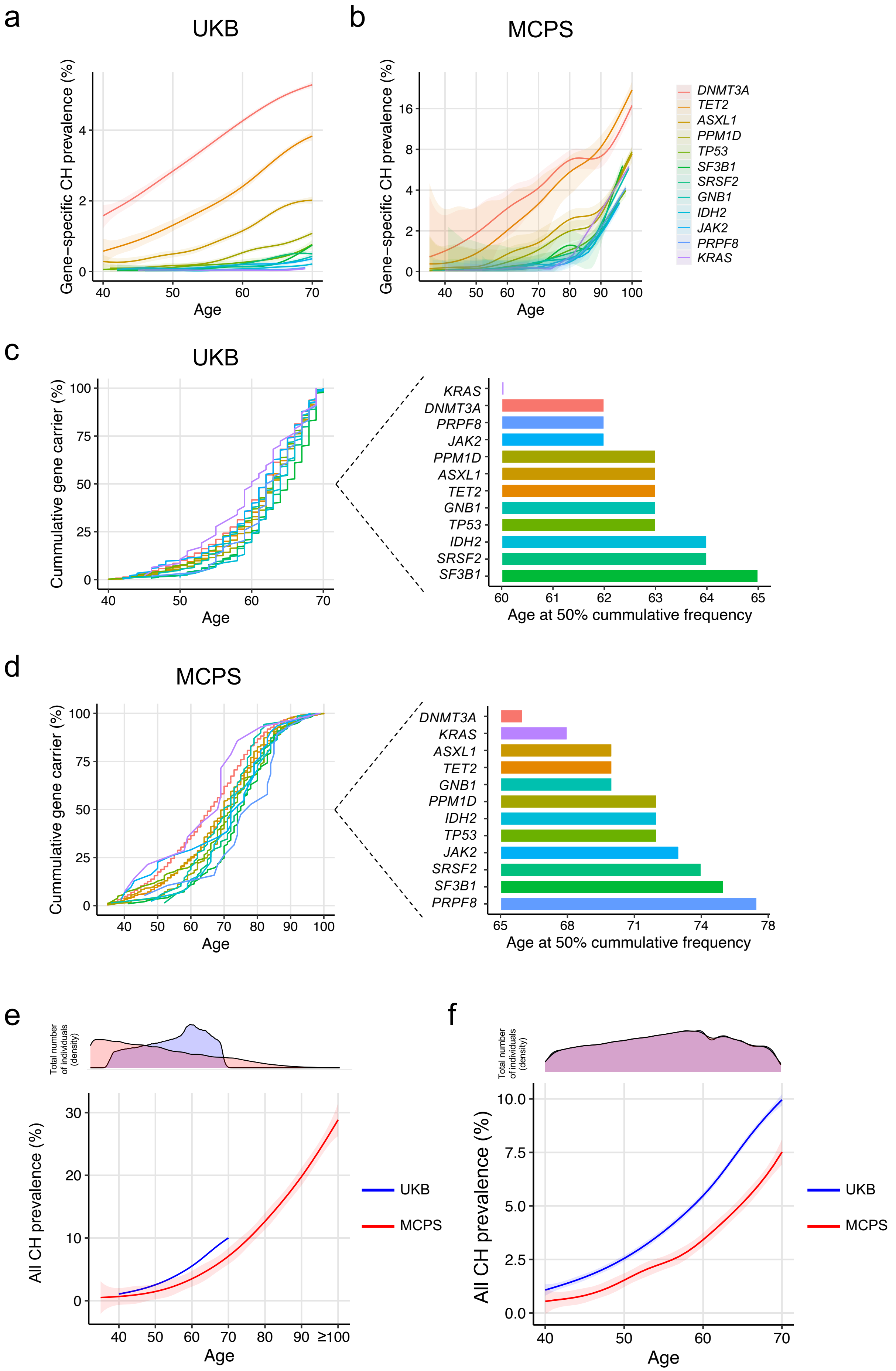

a
UKB
Gene−specific CH prevalence (%)
Age
b
MCPS
Gene−specific CH prevalence (%)
Age
DNMT3A
TET2
ASXL1
PPM1D
TP53
SF3B1
SRSF2
GNB1
IDH2
JAK2
PRPF8
KRAS
c
UKB
Cummulative gene carrier (%)
Age
KRAS
DNMT3A
PRPF8
JAK2
PPM1D
ASXL1
TET2
GNB1
TP53
IDH2
SRSF2
SF3B1
Age at 50% cummulative frequency
d
MCPS
Cummulative gene carrier (%)
Age
DNMT3A
KRAS
ASXL1
TET2
GNB1
PPM1D
IDH2
TP53
JAK2
SRSF2
SF3B1
PRPF8
Age at 50% cummulative frequency
e
Total number of individuals (density)
All CH prevalence (%)
Age
UKB
MCPS
f
Total number of individuals (density)
All CH prevalence (%)
Age
UKB
MCPS

**Extended Data Fig. 2 | Prevalence of CH by age. a,b**, Prevalence of CH individuals by age stratified by gene-specific CH in UKB (**a**) and MCPS (**b**). The centre line represents the fitted values from the general additive model with P-spline smooth class, and the shaded region represents the 95% confidence interval of the fitted values. Cumulative prevalence of CH individuals by age stratified by gene-specific CH in UKB (**c**) and MCPS (**d**). **e**, Same as Figure 1b, but with UKB and MCPS age distribution indicated. **f**, Prevalence of CH individuals by age after age- and sex-matching UKB and MCPS individuals. Overall CH prevalence was 4.55% and 2.84% for UKB and MCPS participants, respectively ($P < 2.2 \times 10^{-16}$). The centre line represents the fitted values from the general additive model with P-spline smooth class, and the shaded region represents the 95% confidence interval of the fitted values.

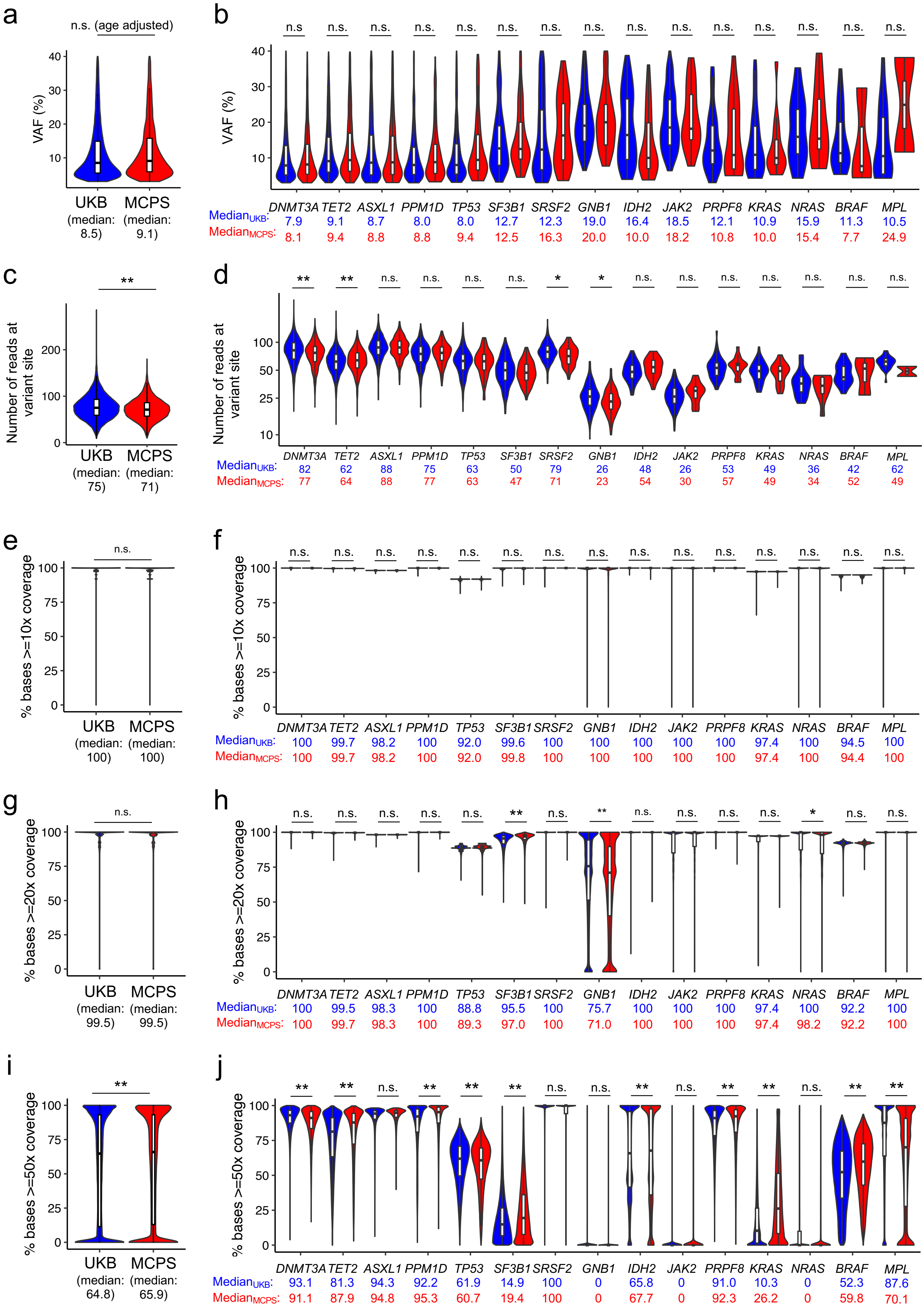
a
n.s. (age adjusted)
VAF (%)
UKB (median: 8.5)
MCPS (median: 9.1)
b
VAF (%)
DNMT3A TET2 ASXL1 PPM1D TP53 SF3B1 SRSF2 GNB1 IDH2 JAK2 PRPF8 KRAS NRAS BRAF MPL
Median_UKB: 7.9 9.1 8.7 8.0 8.0 12.7 12.3 19.0 16.4 18.5 12.1 10.9 15.9 11.3 10.5
Median_MCPS: 8.1 9.4 8.8 8.8 9.4 12.5 16.3 20.0 10.0 18.2 10.8 10.0 15.4 7.7 24.9
c
Number of reads at variant site
UKB (median: 75)
MCPS (median: 71)
d
Number of reads at variant site
DNMT3A TET2 ASXL1 PPM1D TP53 SF3B1 SRSF2 GNB1 IDH2 JAK2 PRPF8 KRAS NRAS BRAF MPL
Median_UKB: 82 62 88 75 63 50 79 26 48 26 53 49 36 42 62
Median_MCPS: 77 64 88 77 63 47 71 23 54 30 57 49 34 52 49
e
% bases >=10x coverage
UKB (median: 100)
MCPS (median: 100)
f
% bases >=10x coverage
DNMT3A TET2 ASXL1 PPM1D TP53 SF3B1 SRSF2 GNB1 IDH2 JAK2 PRPF8 KRAS NRAS BRAF MPL
Median_UKB: 100 99.7 98.2 100 92.0 99.6 100 100 100 100 100 97.4 100 94.5 100
Median_MCPS: 100 99.7 98.2 100 92.0 99.8 100 100 100 100 100 97.4 100 94.4 100
g
% bases >=20x coverage
UKB (median: 99.5)
MCPS (median: 99.5)
h
% bases >=20x coverage
DNMT3A TET2 ASXL1 PPM1D TP53 SF3B1 SRSF2 GNB1 IDH2 JAK2 PRPF8 KRAS NRAS BRAF MPL
Median_UKB: 100 99.5 98.3 100 88.8 95.5 100 75.7 100 100 100 97.4 100 92.2 100
Median_MCPS: 100 99.7 98.3 100 89.3 97.0 100 71.0 100 100 100 97.4 98.2 92.2 100
i
% bases >=50x coverage
UKB (median: 64.8)
MCPS (median: 65.9)
j
% bases >=50x coverage
DNMT3A TET2 ASXL1 PPM1D TP53 SF3B1 SRSF2 GNB1 IDH2 JAK2 PRPF8 KRAS NRAS BRAF MPL
Median_UKB: 93.1 81.3 94.3 92.2 61.9 14.9 100 0 65.8 0 91.0 10.3 0 52.3 87.6
Median_MCPS: 91.1 87.9 94.8 95.3 60.7 19.4 100 0 67.7 0 92.3 26.2 0 59.8 70.1

**Extended Data Fig. 3 | Sequencing coverage comparison of CH genes between UKB and MCPS. a,b**, Comparison of VAF between UKB and MCPS for all CH driver gene variants (**a**) and for gene-specific CH driver gene variants. **c,d**, Number of sequencing reads at site of variants for all CH driver gene variants (**c**) and for gene-specific CH driver gene variants (**d**). **e-j**, Percentage of bases with at least 10x (**e-f**), 20x (**g-h**), and 50x coverage (**i,j**) across all targeted regions of CH driver genes (**e,g,i**) and specific CH driver genes (**f,h,j**). Because VAF is, in part, affected by age, comparison of VAF for overall CH between UKB and MCPS was performed using linear regression with age included as a co-variate, and the resulting age-adjusted *P* value reported in Fig. 3a. Sequencing coverage compared using Wilcoxon ranked sum test. *P* value ** < 0.01 * < 0.05 for overall CH. FDR ** < 0.01 * < 0.05 for gene-specific CH. VAF, variant allele frequency.

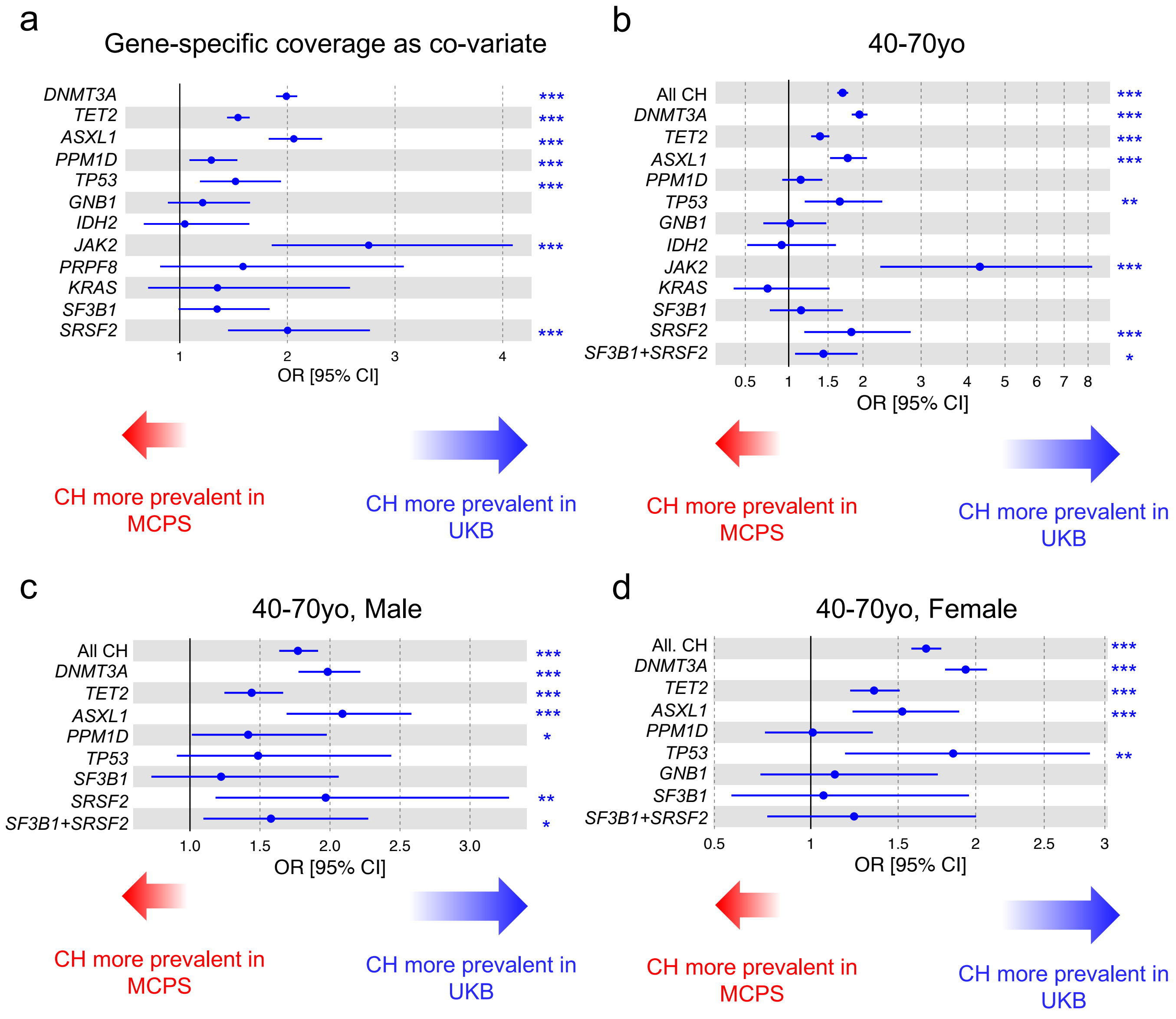

a
Gene-specific coverage as co-variate
DNMT3A ***
TET2 ***
ASXL1 ***
PPM1D ***
TP53 ***
GNB1
IDH2
JAK2 ***
PRPF8
KRAS
SF3B1
SRSF2 ***
1 2 3 4
OR [95% CI]
CH more prevalent in MCPS
CH more prevalent in UKB
b
40-70yo
All CH ***
DNMT3A ***
TET2 ***
ASXL1 ***
PPM1D
TP53 **
GNB1
IDH2
JAK2 ***
KRAS
SF3B1
SRSF2 ***
SF3B1+SRSF2 *
0.5 1 1.5 2 3 4 5 6 7 8
OR [95% CI]
CH more prevalent in MCPS
CH more prevalent in UKB
c
40-70yo, Male
All CH ***
DNMT3A ***
TET2 ***
ASXL1 ***
PPM1D *
TP53
SF3B1
SRSF2 **
SF3B1+SRSF2 *
1.0 1.5 2.0 2.5 3.0
OR [95% CI]
CH more prevalent in MCPS
CH more prevalent in UKB
d
40-70yo, Female
All. CH ***
DNMT3A ***
TET2 ***
ASXL1 ***
PPM1D
TP53 **
GNB1
SF3B1
SF3B1+SRSF2
0.5 1 1.5 2 2.5 3
OR [95% CI]
CH more prevalent in MCPS
CH more prevalent in UKB

**Extended Data Fig. 4 | Inter-population analysis of all CH and gene-specific CH between UKB and MCPS. a,** Comparison of CH prevalence between 419,228 UKB individuals and 136,401 MCPS individuals. Logistic regression model was adjusted for age, sex, and smoking status, and sequencing coverage. **b**, Comparison of CH prevalence between 419,219 UKB individuals and 95,294 MCPS individuals aged 40-70 years of age. Logistic regression model was adjusted for age, sex, and smoking status. **c**, Comparison of CH prevalence between 193,196 UKB males and 31,074 MCPS males aged 40-70 years of age. Logistic regression model was adjusted for age and smoking status. **d**, Comparison of CH prevalence between 226,023 UKB females and 64,220 MCPS females aged 40-70 years of age. Logistic regression model was adjusted for age and smoking status. Only gene-specific CH genes identified in at least 10 individuals were included for analysis. Odds ratio and *P* values were derived from logistic regression model with all CH or gene-specific CH as outcome. yo, years old. *P* value *** < 0.001 ** < 0.01 * < 0.05.

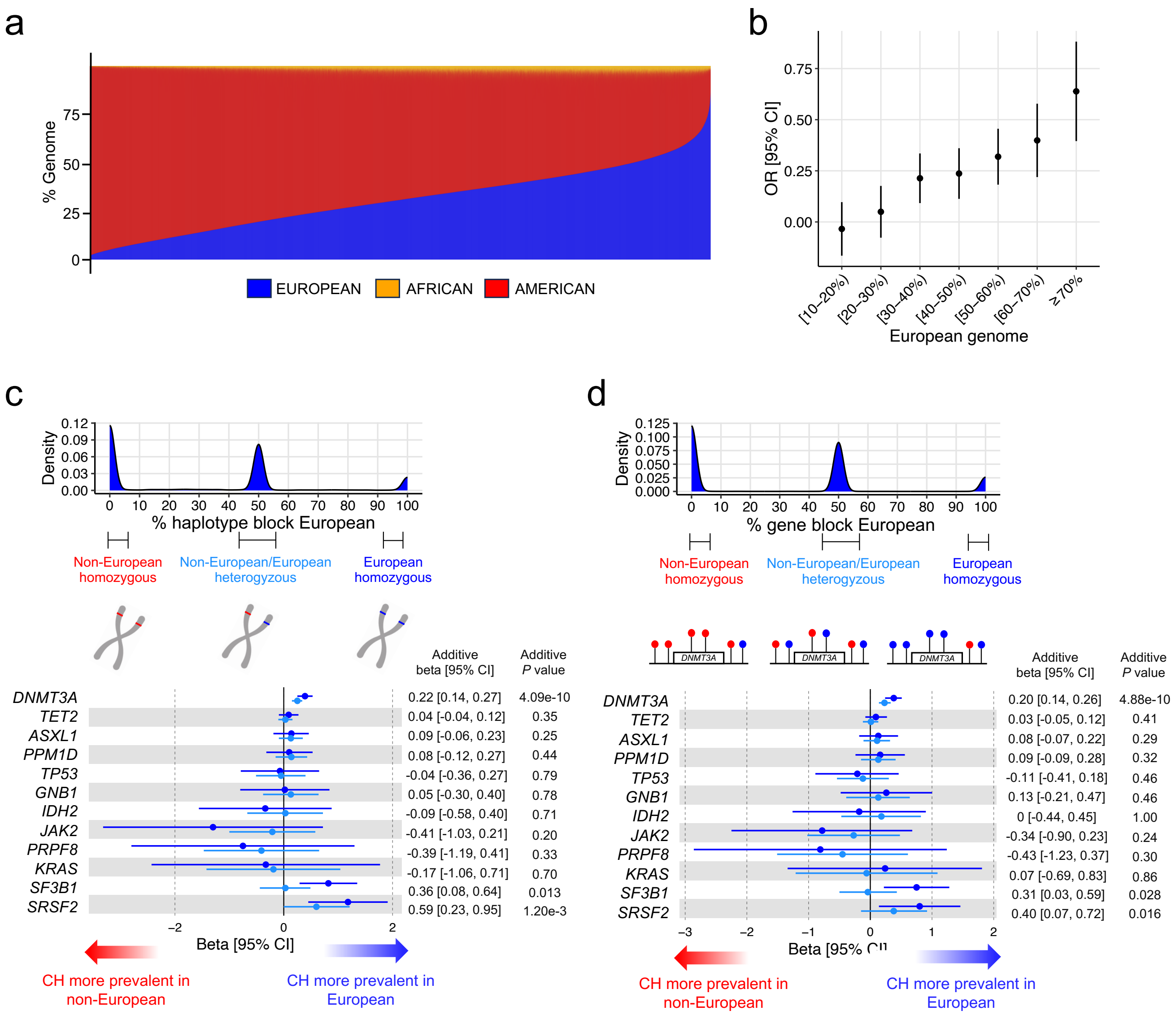
a
% Genome
EUROPEAN
AFRICAN
AMERICAN
b
OR [95% CI]
European genome
c
Density
% haplotype block European
Non-European homozygous
Non-European/European heterogyzous
European homozygous
Additive beta [95% CI]
Additive P value
DNMT3A 0.22 [0.14, 0.27] 4.09e-10
TET2 0.04 [-0.04, 0.12] 0.35
ASXL1 0.09 [-0.06, 0.23] 0.25
PPM1D 0.08 [-0.12, 0.27] 0.44
TP53 -0.04 [-0.36, 0.27] 0.79
GNB1 0.05 [-0.30, 0.40] 0.78
IDH2 -0.09 [-0.58, 0.40] 0.71
JAK2 -0.41 [-1.03, 0.21] 0.20
PRPF8 -0.39 [-1.19, 0.41] 0.33
KRAS -0.17 [-1.06, 0.71] 0.70
SF3B1 0.36 [0.08, 0.64] 0.013
SRSF2 0.59 [0.23, 0.95] 1.20e-3
Beta [95% CI]
CH more prevalent in non-European
CH more prevalent in European
d
Density
% gene block European
Non-European homozygous
Non-European/European heterogyzous
European homozygous
DNMT3A
Additive beta [95% CI]
Additive P value
DNMT3A 0.20 [0.14, 0.26] 4.88e-10
TET2 0.03 [-0.05, 0.12] 0.41
ASXL1 0.08 [-0.07, 0.22] 0.29
PPM1D 0.09 [-0.09, 0.28] 0.32
TP53 -0.11 [-0.41, 0.18] 0.46
GNB1 0.13 [-0.21, 0.47] 0.46
IDH2 0 [-0.44, 0.45] 1.00
JAK2 -0.34 [-0.90, 0.23] 0.24
PRPF8 -0.43 [-1.23, 0.37] 0.30
KRAS 0.07 [-0.69, 0.83] 0.86
SF3B1 0.31 [0.03, 0.59] 0.028
SRSF2 0.40 [0.07, 0.72] 0.016
Beta [95% CI]
CH more prevalent in non-European
CH more prevalent in European

**Extended Data Fig. 5 | Intra-population comparison of the prevalence of all CH and gene-specific CH in MCPS.** **a**, On the y-axis is the proportion of European, Indigenous American, and African genome across the MCPS individuals on the x-axis. **b**, Overall CH risk associated with binned proportion of European genome relative to individuals with 0-10% European genome adjusted for age, sex, and smoking status. **c,d**, Prevalence of gene-specific CH in MCPS individuals with European (homozygous or heterozygous) vs non-European (homozygous American or African) haplotype block (**c**) or genomic block (**d**) in which the corresponding CH driver gene is located. Beta coefficients and *P* values were derived from logistic regression model with all CH or gene-specific CH as outcome, and with age, sex, and smoking status included as co-variates.

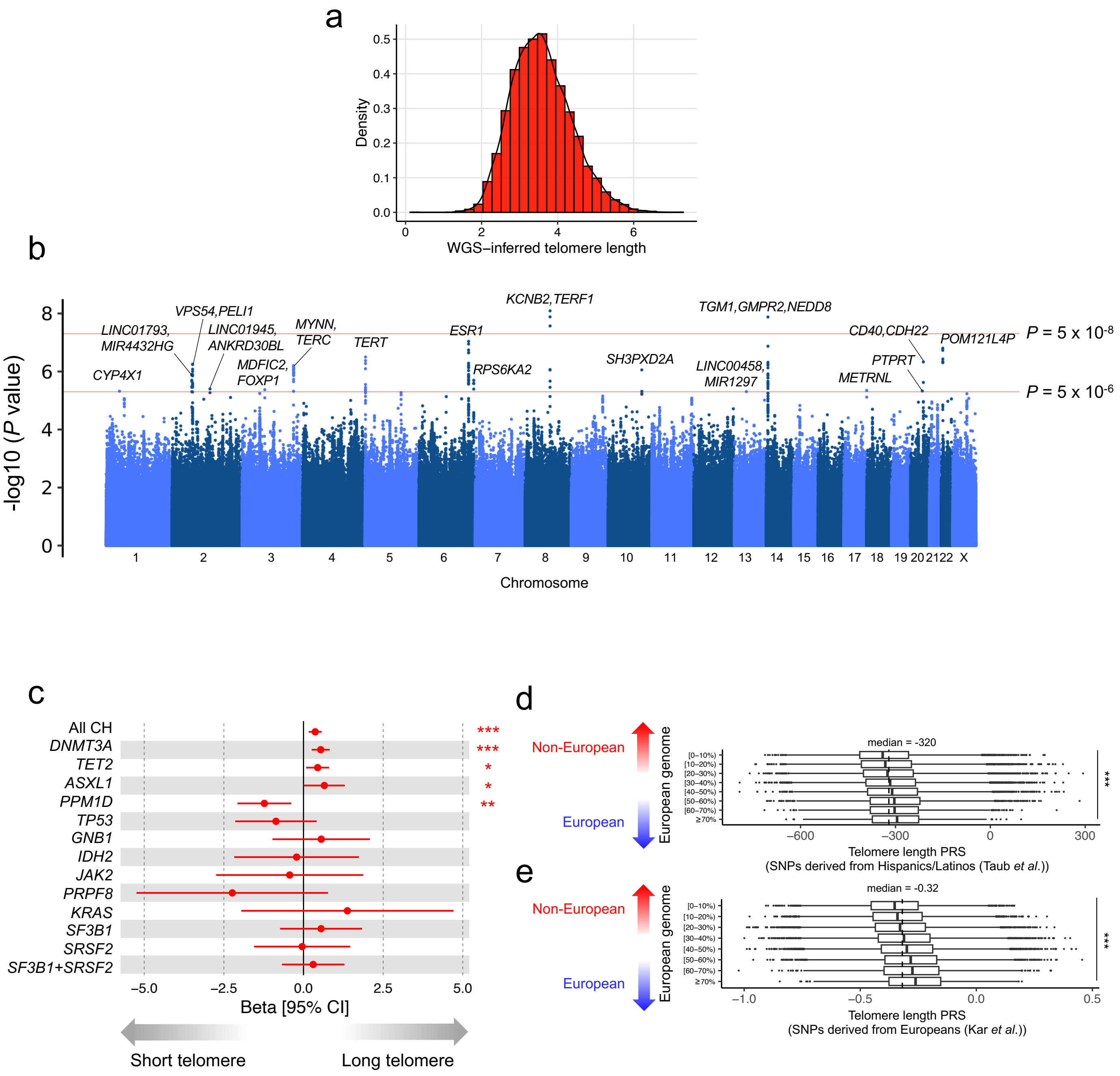

a
Density
WGS–inferred telomere length
b
-log10 (P value)
Chromosome
CYP4X1
LINC01793, MIR4432HG
VPS54,PELI1
LINC01945, ANKRD30BL
MDFIC2, FOXP1
MYNN, TERC
TERT
ESR1
RPS6KA2
KCNB2,TERF1
SH3PXD2A
TGM1,GMPR2,NEDD8
LINC00458, MIR1297
METRNL
PTPRT
CD40,CDH22
POM121L4P
P = 5 x 10-8
P = 5 x 10-6
c
All CH
DNMT3A
TET2
ASXL1
PPM1D
TP53
GNB1
IDH2
JAK2
PRPF8
KRAS
SF3B1
SRSF2
SF3B1+SRSF2
Beta [95% CI]
Short telomere
Long telomere
d
Non-European
European
European genome
median = -320
Telomere length PRS
(SNPs derived from Hispanics/Latinos (Taub et al.))
e
Non-European
European
European genome
median = -0.32
Telomere length PRS
(SNPs derived from Europeans (Kar et al.))

**Extended Data Fig. 6 | Telomere length GWAS and PRS in MCPS. a,** Distribution of WGS-inferred telemore length across 9,469 individuals with WGS data available. **b**, Manhattan plot of telomere length GWAS results. *P*-values on y-axis were derived from Firth logistic regression implemented by REGENIE software. **c**, Association between PRS-predicted telomere length and overall CH and gene-specific CH. Beta coefficients and *P* values were derived from logistic regression model with all CH or gene-specific CH as outcome, and with age, sex, and smoking status included as co-variates. **d,e**, Distribution of PRS-predicted telomere length across individuals with varying degree of European genome. PRS was built using telomere length-associated SNPs derived from previously reported GWAS analysis of Hispanics/Latinos from TOPMED (d) and of Europeans from UKB (e). *P*-values were derived from Kruskal-Wallis test. PRS, polygenic risk score. *P* value *** < 0.001 ** < 0.01 * < 0.05.

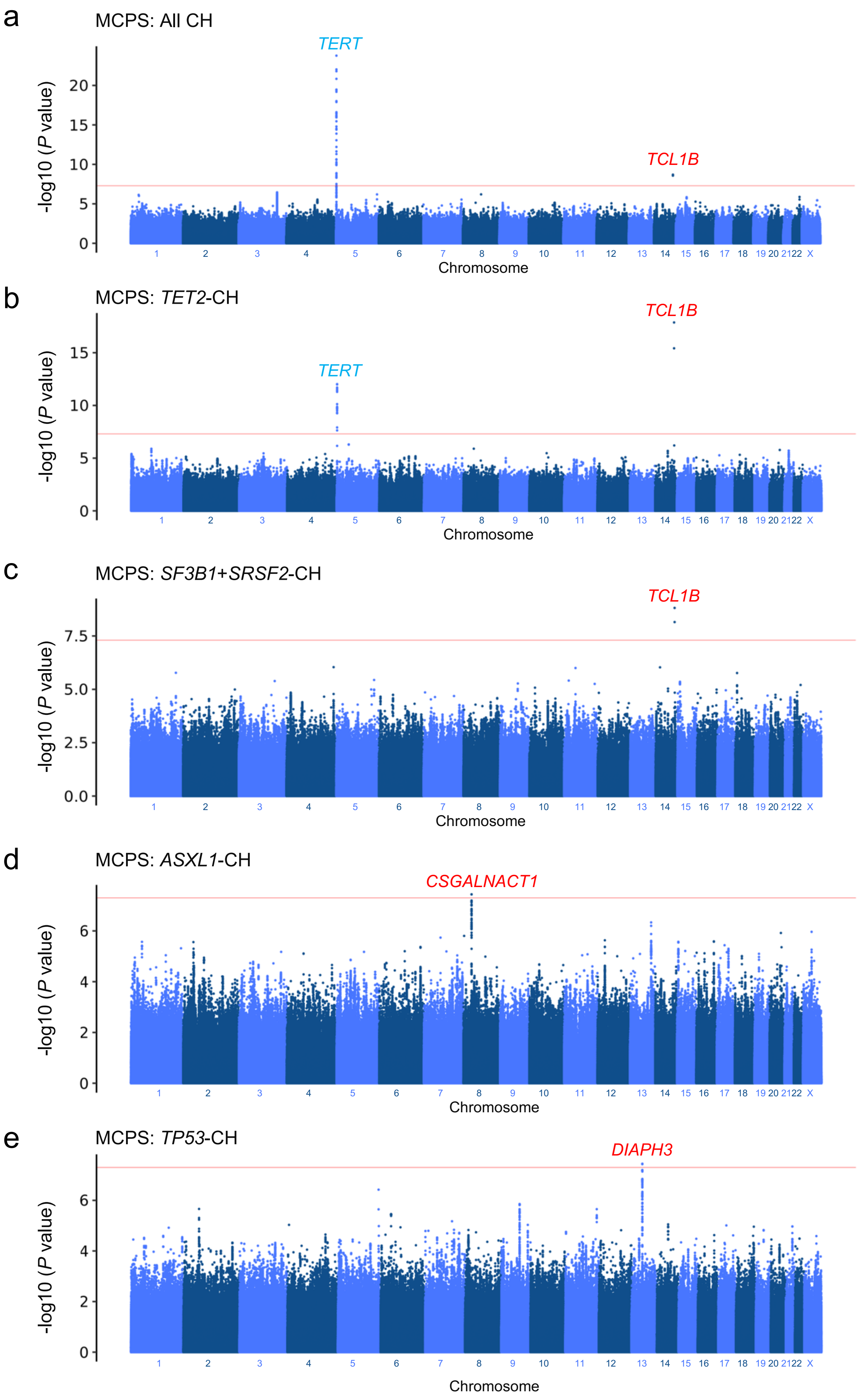
a
MCPS: All CH
TERT
TCL1B
-log10 (P value)
0
5
10
15
20
Chromosome
1 2 3 4 5 6 7 8 9 10 11 12 13 14 15 16 17 18 19 20 21 22 X
b
MCPS: TET2-CH
TCL1B
TERT
-log10 (P value)
0
5
10
15
Chromosome
c
MCPS: SF3B1+SRSF2-CH
TCL1B
-log10 (P value)
0.0
2.5
5.0
7.5
Chromosome
d
MCPS: ASXL1-CH
CSGALNACT1
-log10 (P value)
0
2
4
6
Chromosome
e
MCPS: TP53-CH
DIAPH3
-log10 (P value)
0
2
4
6
Chromosome

**Extended Data Fig. 7 | GWAS of all CH and gene-specific CH in MCPS. a-e**, Manhattan plot representing the common genetic variants (MAF ≥ 1%) included for GWAS in MCPS for overall CH (**a**), and *TET2*- (**b**), splicing factor- (**c**), *ASXL1*- (**d**), and *TP53*- (**e**) CH. *P* values on y-axis were derived from Firth logistic regression implemented by REGENIE software. Three novel signals (red) were identified as genome-wide significant (*P* value < $5 \times 10^{-8}$, red horizontal line) with the nearest gene of the leading SNP annotated for the respective locus. Previously reported associations from European populations indicated in blue.

**a**

| | | rs10131341 (*TCL1A* upstream) | |
|---|---|---|---|
| | | A | C |
| rs187319135 (*TCL1B* upstream) | C | 78.29% | 20.72% |
| | T | 0.98% | 0.01% |

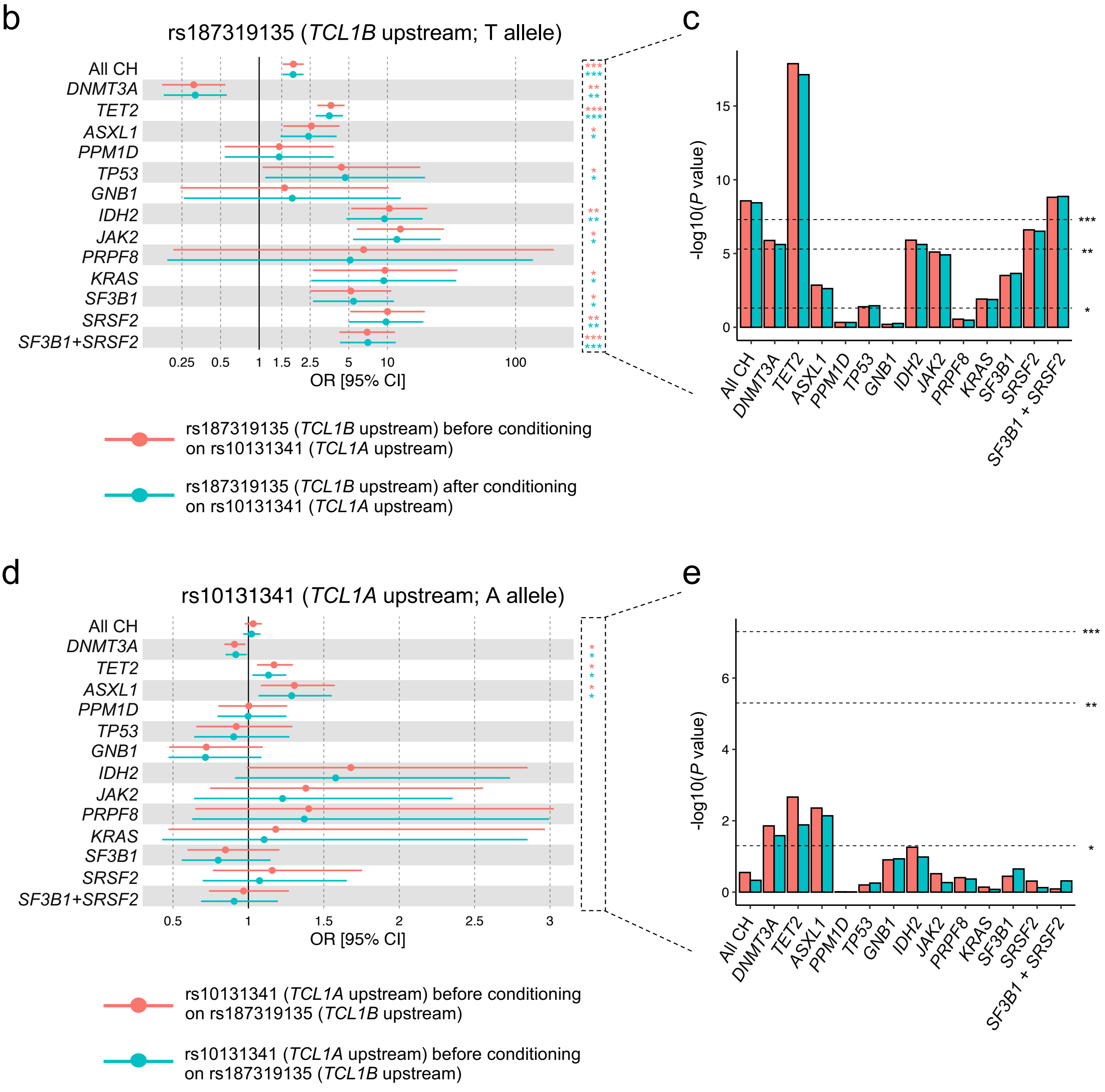

**Extended Data Fig. 8 | Linkage and conditional analysis of rs187319135 (TCL1B upstream) and rs10131341 (TCL1A upstream) variants in MCPS. a,** Phasing of rs187319135 and rs10131341 using PLINK2. These variants are in high LD with each other (D' = 0.95, $r^2$ = 0.024). **b,c**, Risk conferred by rs187319135 (T allele) to overall CH and gene-specific CH before and after conditioning on rs10131341. **d,e**, Risk conferred by rs10131341 (A allele) to overall CH and gene-specific CH before and after conditioning on rs187319135.

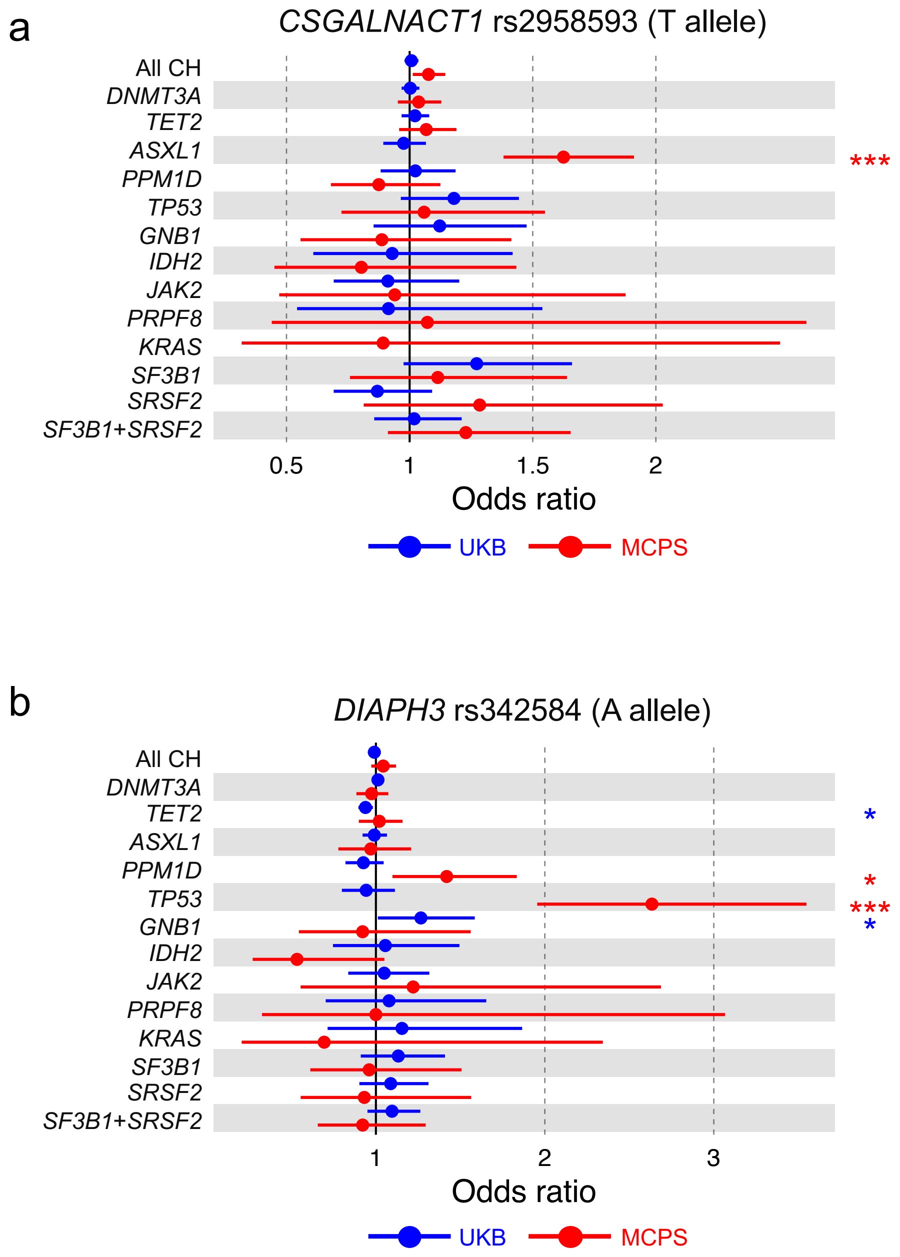
a
CSGALNACT1 rs2958593 (T allele)
All CH
DNMT3A
TET2
ASXL1
PPM1D
TP53
GNB1
IDH2
JAK2
PRPF8
KRAS
SF3B1
SRSF2
SF3B1+SRSF2
***
0.5
1
1.5
2
Odds ratio
UKB
MCPS
b
DIAPH3 rs342584 (A allele)
All CH
DNMT3A
TET2
ASXL1
PPM1D
TP53
GNB1
IDH2
JAK2
PRPF8
KRAS
SF3B1
SRSF2
SF3B1+SRSF2
*
*
***
*
1
2
3
Odds ratio
UKB
MCPS

**Extended Data Fig. 9 | Risk estimates of novel common CH risk variants identified from GWAS in MCPS. a**, rs2958593 identified as genome-wide significant from *ASXL1*-CH. Overall CH and gene-specific CH risk estimates conferred by the minor allele (T) shown here. **b**, rs342584 identified as genome-wide significant from *TP53*-CH. Overall CH and gene-specific CH risk estimates conferred by the minor allele (A) shown here. *P* value *** < $5 \times 10^{-8}$ (genome-wide significant) ** < $5 \times 10^{-6}$ (suggestive) * < 0.05 (nominal).

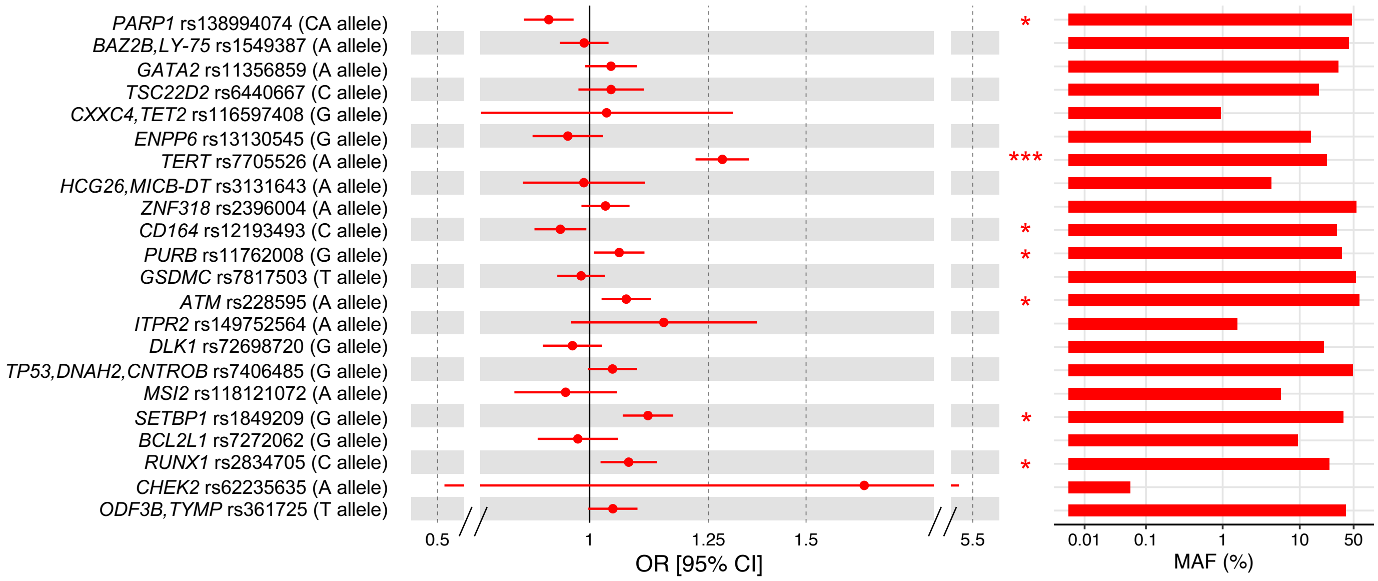
PARP1 rs138994074 (CA allele)
BAZ2B,LY-75 rs1549387 (A allele)
GATA2 rs11356859 (A allele)
TSC22D2 rs6440667 (C allele)
CXXC4,TET2 rs116597408 (G allele)
ENPP6 rs13130545 (G allele)
TERT rs7705526 (A allele)
HCG26,MICB-DT rs3131643 (A allele)
ZNF318 rs2396004 (A allele)
CD164 rs12193493 (C allele)
PURB rs11762008 (G allele)
GSDMC rs7817503 (T allele)
ATM rs228595 (A allele)
ITPR2 rs149752564 (A allele)
DLK1 rs72698720 (G allele)
TP53,DNAH2,CNTROB rs7406485 (G allele)
MSI2 rs118121072 (A allele)
SETBP1 rs1849209 (G allele)
BCL2L1 rs7272062 (G allele)
RUNX1 rs2834705 (C allele)
CHEK2 rs62235635 (A allele)
ODF3B,TYMP rs361725 (T allele)
0.5
1
1.25
1.5
5.5
OR [95% CI]
*
***
*
*
*
*
*
0.01
0.1
1
10
50
MAF (%)

**Extended Data Fig. 10 | Summary statistics of MCPS GWAS of overall CH for common risk variants previously identified from European populations.** Risk estimates conferred by reported risk variants and MAF for the respective risk variants estimated from our study indicated. Note that previously reported leading SNPs for *TRIM59/IFT80* (rs56658671) and *STN1* (rs34763036) loci were not detected in MCPS imputed genetic data, and hence not included for replication assessment here. MAF, minor allele frequency; OR, odds ratio. *P* value *** < 5 x $10^{-8}$ (genome-wide significant) ** < 5 x $10^{-6}$ (suggestive) * < 0.05 (nominal).

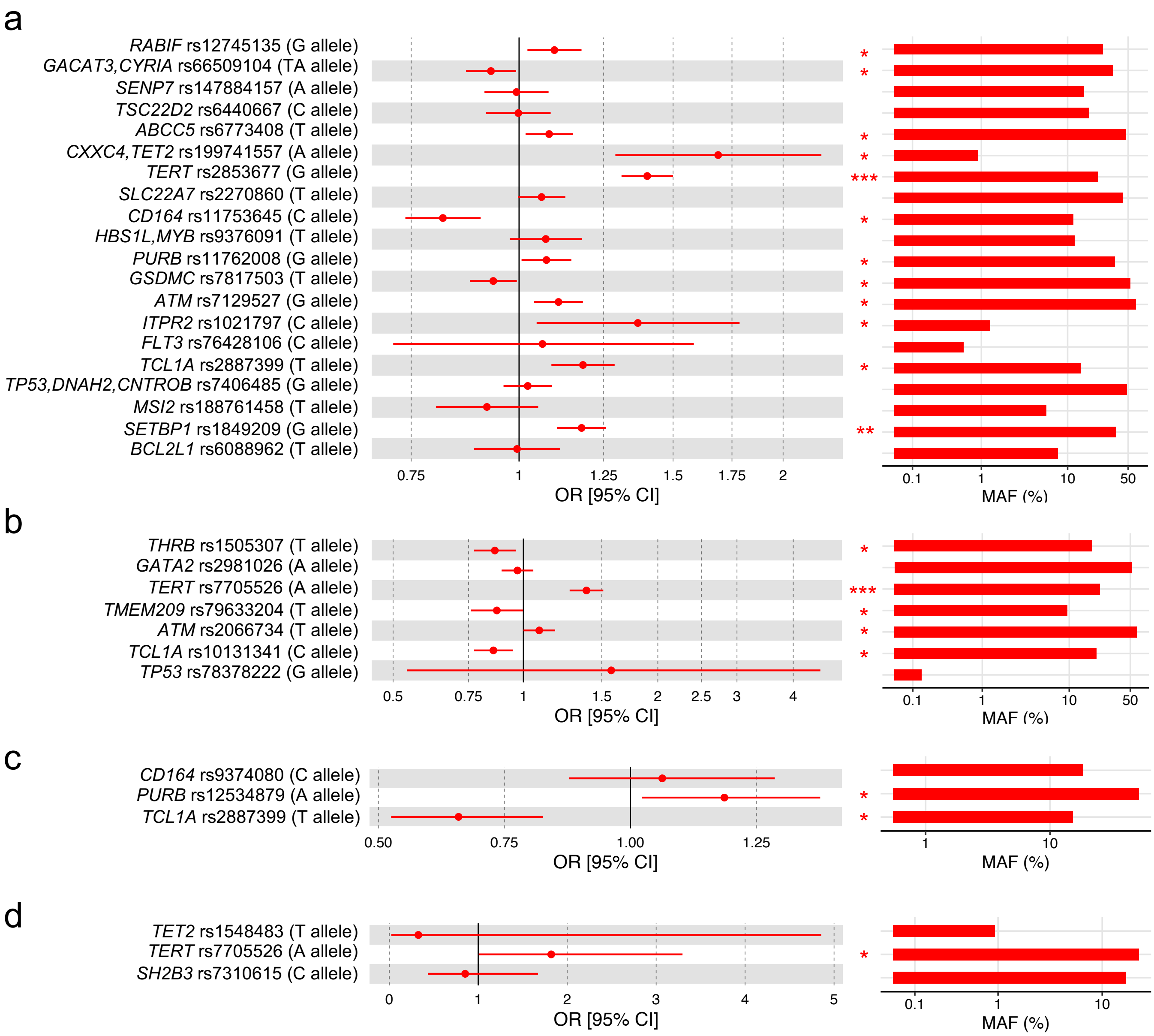
a
RABIF rs12745135 (G allele)
GACAT3,CYRIA rs66509104 (TA allele)
SENP7 rs147884157 (A allele)
TSC22D2 rs6440667 (C allele)
ABCC5 rs6773408 (T allele)
CXXC4,TET2 rs199741557 (A allele)
TERT rs2853677 (G allele)
SLC22A7 rs2270860 (T allele)
CD164 rs11753645 (C allele)
HBS1L,MYB rs9376091 (T allele)
PURB rs11762008 (G allele)
GSDMC rs7817503 (T allele)
ATM rs7129527 (G allele)
ITPR2 rs1021797 (C allele)
FLT3 rs76428106 (C allele)
TCL1A rs2887399 (T allele)
TP53,DNAH2,CNTROB rs7406485 (G allele)
MSI2 rs188761458 (T allele)
SETBP1 rs1849209 (G allele)
BCL2L1 rs6088962 (T allele)
0.75
1
1.25
1.5
1.75
2
OR [95% CI]
0.1
1
10
50
MAF (%)
b
THRB rs1505307 (T allele)
GATA2 rs2981026 (A allele)
TERT rs7705526 (A allele)
TMEM209 rs79633204 (T allele)
ATM rs2066734 (T allele)
TCL1A rs10131341 (C allele)
TP53 rs78378222 (G allele)
0.5
0.75
1
1.5
2
2.5
3
4
OR [95% CI]
0.1
1
10
50
MAF (%)
c
CD164 rs9374080 (C allele)
PURB rs12534879 (A allele)
TCL1A rs2887399 (T allele)
0.50
0.75
1.00
1.25
OR [95% CI]
1
10
MAF (%)
d
TET2 rs1548483 (T allele)
TERT rs7705526 (A allele)
SH2B3 rs7310615 (C allele)
0
1
2
3
4
5
OR [95% CI]
0.1
1
10
MAF (%)

**Extended Data Fig. 11 | Summary statistics of MCPS GWAS of gene-specific CH for common risk variants previously identified from European populations. a-d**, Risk estimates conferred by reported risk variants for *DNMT3A*- (**a**), *TET2*- (**b**), *ASXL1*- (**c**), and *JAK2*- (**d**) CH. The MAF for the respective risk variants estimated from our study also indicated. Note that previously reported leading SNPs for *PARP1* (rs118165556), *PLA2R1* (rs777836095), *TRIM59/IFT80* (rs11418097), and *STN1* (rs34763036) loci for *DNMT3A*-CH, and *JAK2* (rs574643736) locus for *JAK2*-CH were not detected in MCPS imputed genetic data, and hence not included for replication assessment here. MAF, minor allele frequency; OR, odds ratio. *P* value *** < $5 \times 10^{-8}$ (genome-wide significant) ** < $5 \times 10^{-6}$ (suggestive) * < 0.05 (nominal).

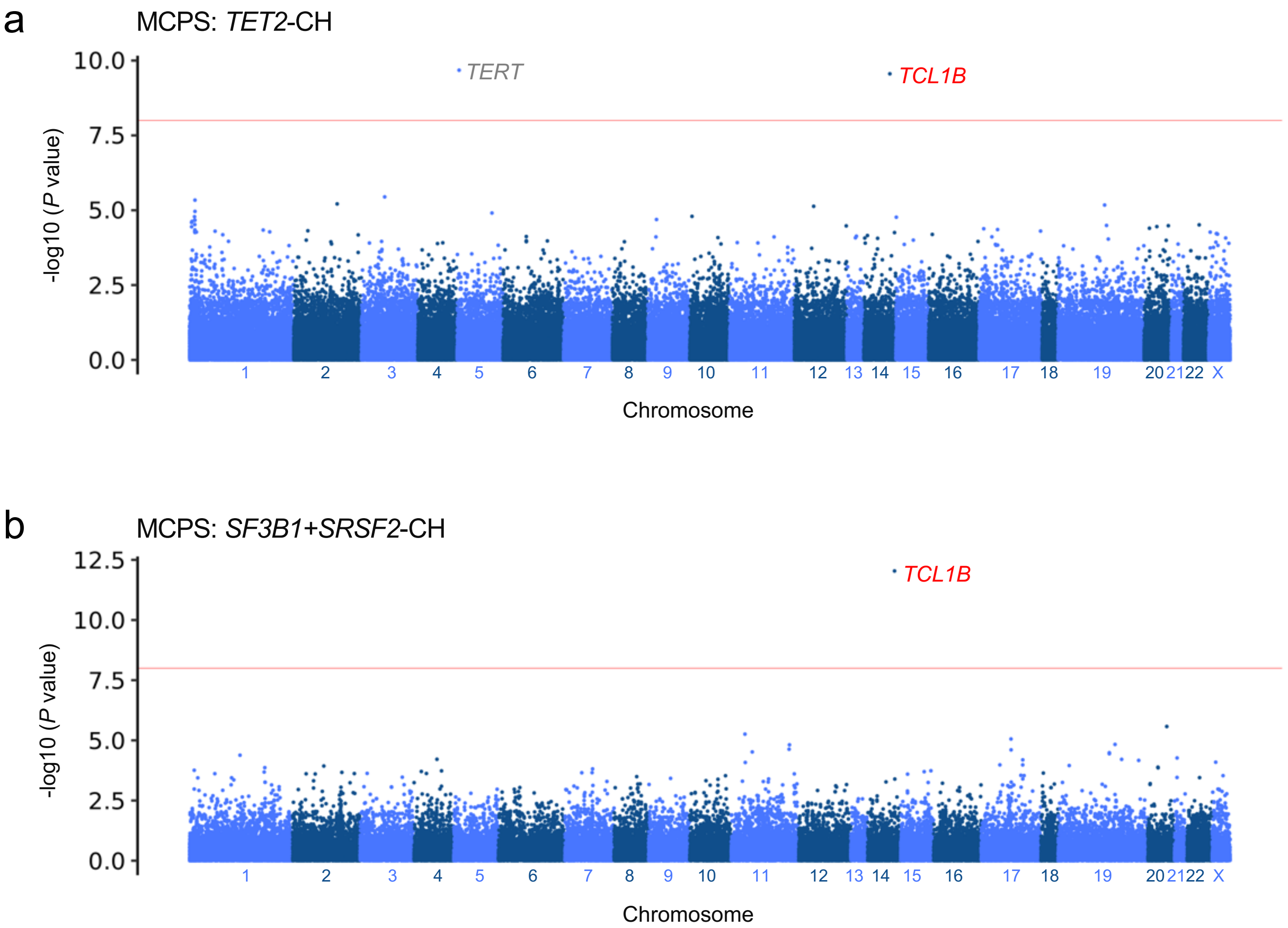
a
MCPS: TET2-CH
TERT
TCL1B
-log10 (P value)
10.0
7.5
5.0
2.5
0.0
X
Chromosome
b
MCPS: SF3B1+SRSF2-CH
TCL1B
-log10 (P value)
12.5
10.0
7.5
5.0
2.5
0.0
Chromosome

**Extended Data Fig. 12 | ExWAS of gene-specific CH in MCPS. a-b**, Manhattan plots representing the common (MAF ≥ 1%) and rare (MAF < 1%) genetic variants included for ExWAS of *TET2*- (**a**) and *SF3B1*+*SRSF2*- (**b**) CH. *P*-values on y-axis were derived from Firth logistic regression implemented by REGENIE software. Previously reported associations from GWAS indicated in grey, and novel associations from ExWAS indicated in red. *P* values on y-axis were derived from Fisher's exact test. Nearest gene of the leading SNP annotated for the respective locus.

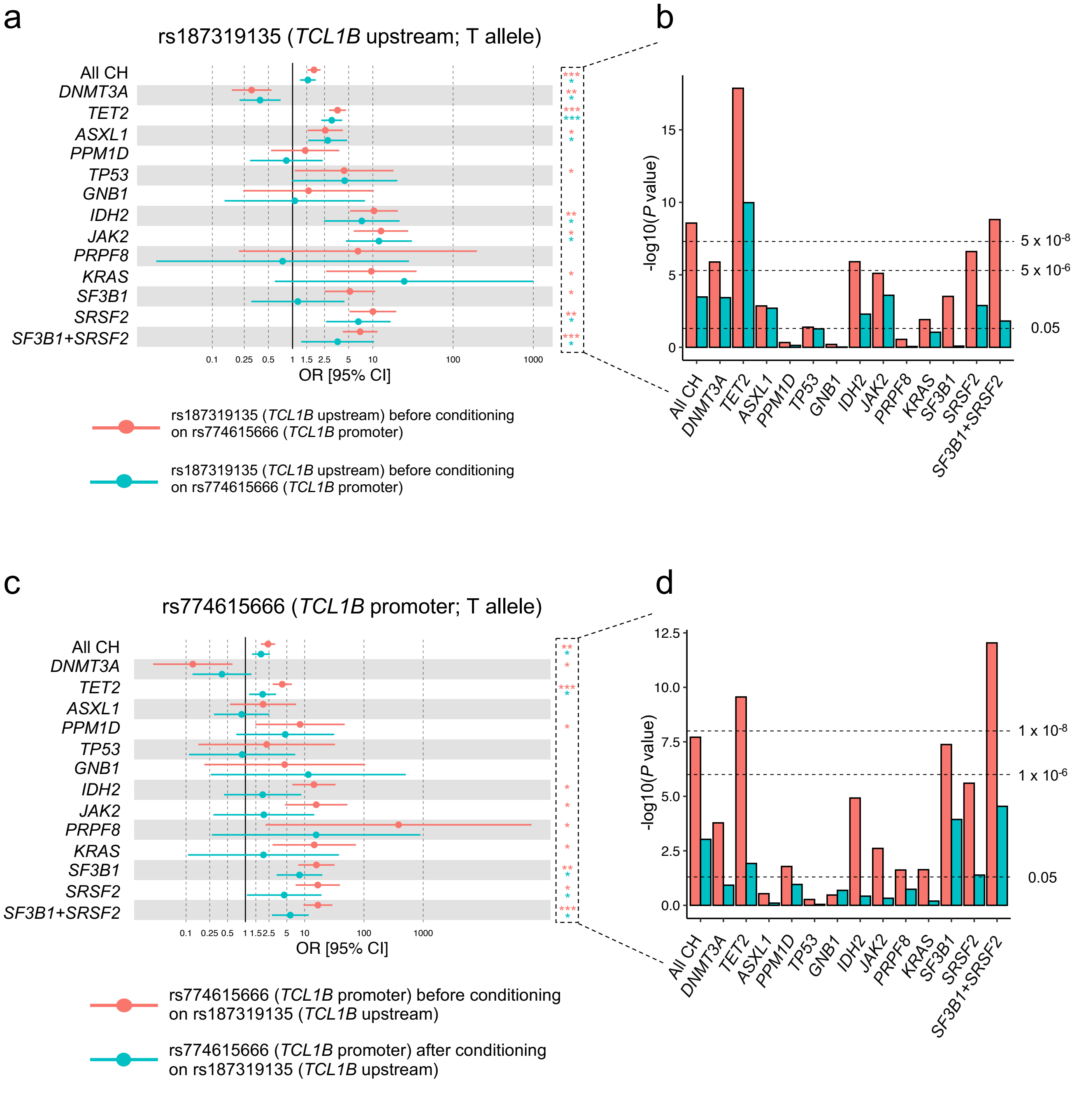

a
rs187319135 (TCL1B upstream; T allele)
All CH
DNMT3A
TET2
ASXL1
PPM1D
TP53
GNB1
IDH2
JAK2
PRPF8
KRAS
SF3B1
SRSF2
SF3B1+SRSF2
0.1
0.25
0.5
1
1.5
2.5
5
10
100
1000
OR [95% CI]
rs187319135 (TCL1B upstream) before conditioning on rs774615666 (TCL1B promoter)
rs187319135 (TCL1B upstream) before conditioning on rs774615666 (TCL1B promoter)
b
-log10(P value)
15
10
5
0
5 x 10-8
5 x 10-6
0.05
c
rs774615666 (TCL1B promoter; T allele)
OR [95% CI]
rs774615666 (TCL1B promoter) before conditioning on rs187319135 (TCL1B upstream)
rs774615666 (TCL1B promoter) after conditioning on rs187319135 (TCL1B upstream)
d
-log10(P value)
12.5
10.0
7.5
5.0
2.5
0.0
1 x 10-8
1 x 10-6
0.05

**Extended Data Fig. 13 | Conditional analysis of rs187319135 (TCL1B upstream) and rs774615666 (TCL1B promoter) variants in MCPS. a,b**, Risk conferred by rs187319135 (T allele) to overall CH and gene-specific CH before and after conditioning on rs774615666. Genotype of rs774615666 was determined from whole-exome sequencing (WES), and included as co-variate in the Firth logistic regression model implemented by REGENIE. **c,d**, Risk conferred by rs774615666 (T allele) to overall CH and gene-specific CH before and after conditioning on rs187319135. Genotype of rs187319135 was hard-called from the imputed genetic data, and included as co-variate in the Firth logistic regression model implemented by REGENIE. Thresholds for hard-calling genotypes were $0 \leq x \leq 0.1$, $0.9 \leq x \leq 1.1$, and $1.9 \leq x \leq 2.0$ for homozygous minor allele, heterozygous minor/major allele, and homozygous major allele, respectively, where $x$ is the allelic dosage (expected number of copies of major allele). Allelic dosages outside the range of thresholds were coded as missing. e, Number of individuals with CH carrying the rs187319135 or rs774615666 risk alleles.

**a**

| | | rs774615666 (*TCL1B* promoter) | | |
|---|---|---|---|---|
| | | C/C | C/T | T/T |
| rs187319135 (*TCL1B* upstream) | C/C | 132,414 | 105 | 0 |
| | C/T | 1,458 | 664 | 0 |
| | T/T | 5 | 2 | 3 |

**b**

| | Number of cases with corresponding genotype | Number of controls with corresponding genotype | *P* value |
|---|---|---|---|
| All CH | 69 | 1394 | 2.53E-04 |
| | 7 | 98 | 6.32E-02 |
| | 49 | 620 | 5.09E-12 |
| *DNMT3A* | 6 | 1394 | 1.91E-03 |
| | 0 | 98 | 9.01E-01 |
| | 1 | 620 | 3.58E-02 |
| *TET2* | 42 | 1394 | 3.95E-13 |
| | 5 | 98 | 1.16E-03 |
| | 25 | 620 | 5.33E-15 |
| *ASXL1* | 11 | 1394 | 1.23E-03 |
| | 0 | 98 | 9.61E-01 |
| | 4 | 620 | 3.95E-02 |
| *PPM1D* | 3 | 1394 | 7.06E-01 |
| | 1 | 98 | 1.54E-01 |
| | 2 | 620 | 2.09E-01 |
| *TP53* | 4 | 1394 | 8.28E-03 |
| | 0 | 98 | 9.76E-01 |
| | 1 | 620 | 3.32E-01 |
| *GNB1* | 0 | 1394 | 9.77E-01 |
| | 0 | 98 | 9.93E-01 |
| | 1 | 620 | 1.83E-01 |
| *IDH2* | 2 | 1394 | 2.51E-02 |
| | 0 | 98 | 9.85E-01 |
| | 4 | 620 | 8.82E-09 |
| *JAK2* | 3 | 1394 | 6.42E-05 |
| | 0 | 98 | 9.87E-01 |
| | 2 | 620 | 2.81E-05 |
| *PRPF8* | 0 | 1394 | 9.90E-01 |
| | 0 | 98 | 9.97E-01 |
| | 1 | 620 | 1.25E-02 |
| *KRAS* | 1 | 1394 | 4.97E-02 |
| | 0 | 98 | 9.92E-01 |
| | 1 | 620 | 5.08E-03 |
| *SF3B1* | 2 | 1394 | 2.86E-01 |
| | 2 | 98 | 9.39E-06 |
| | 6 | 620 | 3.69E-12 |
| *SRSF2* | 5 | 1394 | 3.52E-06 |
| | 1 | 98 | 7.77E-03 |
| | 5 | 620 | 5.20E-12 |
| *SF3B1+SRSF2* | 7 | 1394 | 7.56E-05 |
| | 3 | 98 | 4.89E-07 |
| | 11 | 620 | 1.45E-21 |

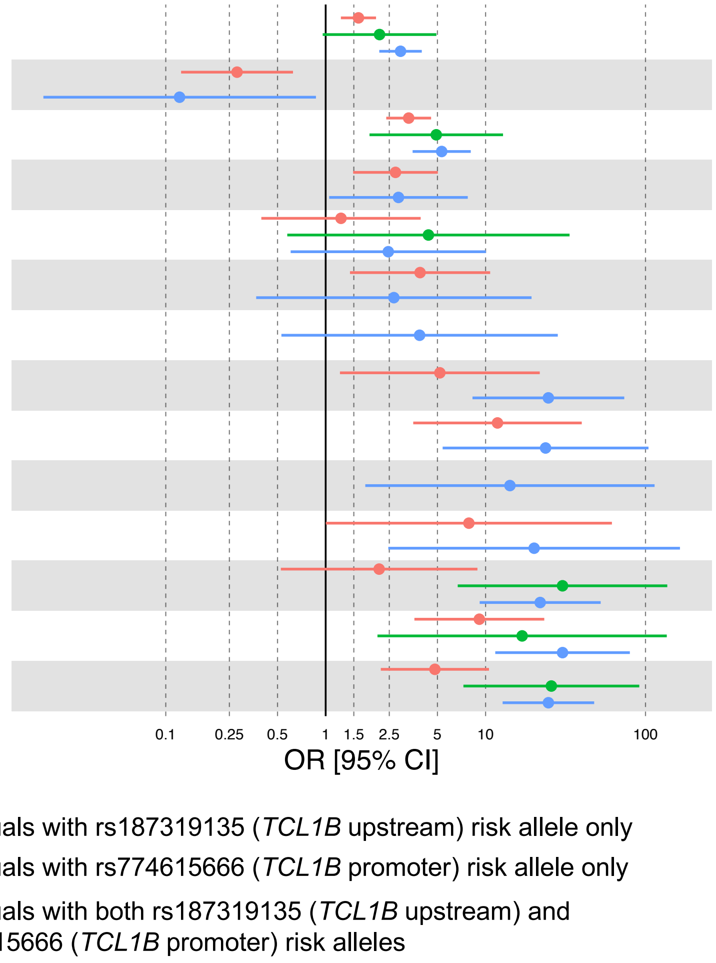

**Extended Data Fig. 14 | Overall CH and gene-specific CH risk conferred by genotypes based on rs187319135 (*TCL1B* upstream) and rs774615666 (*TCL1B* promoter). a,** Cross tabulation of the rs187319135 and rs774615666 genotypes. **b**, Overall CH and gene-specific CH risk estimates in individuals with rs187319135 risk (T) allele only, individuals with rs774615666 risk (T) allele only, and individuals with both rs187319135 and rs774615666 risk alleles relative to individuals with no risk alleles for both rs187319135 and rs774615666.

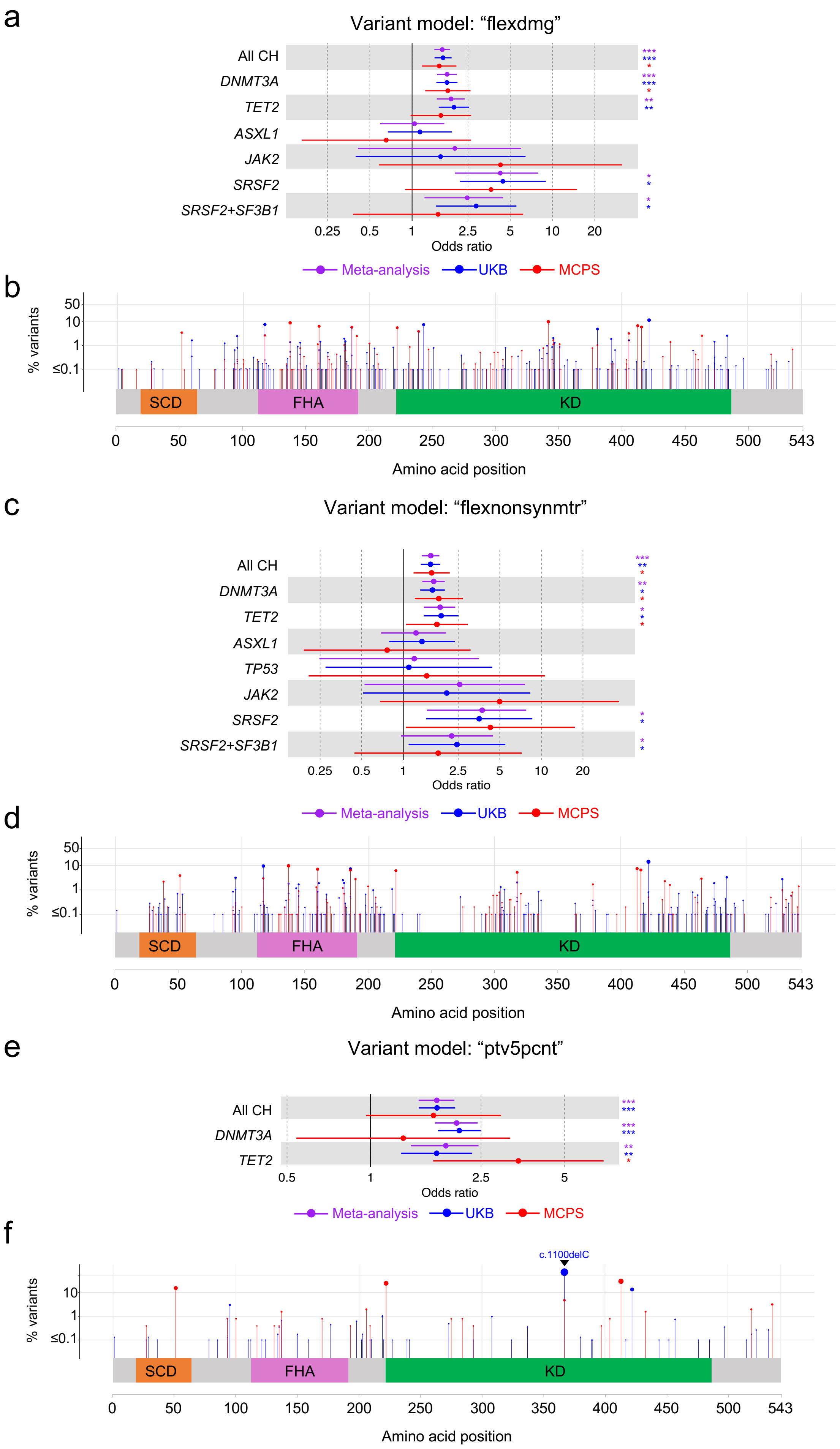
a
Variant model: "flexdmg"
All CH
DNMT3A
TET2
ASXL1
JAK2
SRSF2
SRSF2+SF3B1
0.25 0.5 1 2.5 5 10 20
Odds ratio
Meta-analysis
UKB
MCPS
b
% variants
50
10
1
≤0.1
SCD
FHA
KD
0 50 100 150 200 250 300 350 400 450 500 543
Amino acid position
c
Variant model: "flexnonsynmtr"
All CH
DNMT3A
TET2
ASXL1
TP53
JAK2
SRSF2
SRSF2+SF3B1
0.25 0.5 1 2.5 5 10 20
Odds ratio
Meta-analysis
UKB
MCPS
d
% variants
50
10
1
≤0.1
SCD
FHA
KD
0 50 100 150 200 250 300 350 400 450 500 543
Amino acid position
e
Variant model: "ptv5pcnt"
All CH
DNMT3A
TET2
0.5 1 2.5 5
Odds ratio
Meta-analysis
UKB
MCPS
f
c.1100delC
% variants
10
1
≤0.1
SCD
FHA
KD
0 50 100 150 200 250 300 350 400 450 500 543
Amino acid position

**Extended Data Fig. 15 | Rare *CHEK2* variant burden association meta-analysis across MCPS UKB. a-b**, *CHEK2* “flexdmg” qualifying variant model identified as genome-wide significant (*P* value < 1 x $10^{-8}$) in overall CH and *DNMT3A*-CH. **c-d**, *CHEK2* “flexdnonsynmtr” qualifying variant model identified as genome-wide significant in overall CH. **e-f**, *CHEK2* “ptv5pcnt” qualifying variant model identified as genome-wide significant in overall CH. P values were derived from Cochran-Mantel-Haenszel (CMH) test. For each qualifying variant model, the risk estimates conferred (top) and individual variant as a percentage all *CHEK2* variants identified in MCPS and UKB (bottom) shown. *P* value *** < 1 x $10^{-8}$ ** < 1 x $10^{-6}$ (suggestive) * < 0.05 (nominal).
